# Imported SARS-COV-2 Variants of Concern Drove Spread of Infections Across Kenya During the Second Year of the Pandemic

**DOI:** 10.1101/2022.02.28.22271467

**Authors:** Carolyne Nasimiyu, Damaris Matoke-Muhia, Gilbert K. Rono, Eric Osoro, Daniel O. Obado, J. Milkah Mwangi, Nicholas Mwikwabe, Kelvin Thiong’o, Jeanette Dawa, Isaac Ngere, John Gachohi, Samuel Kariuki, Evans Amukoye, Marianne Mureithi, Philip Ngere, Patrick Amoth, Ian Were, Lyndah Makayotto, Vishvanath Nene, Edward O. Abworo, M. Kariuki Njenga, Stephanie N. Seifert, Samuel O. Oyola

**Author notes:** Joint senior and corresponding authors **CORRESPONDING AUTHORS:** M. Kariuki Njenga, Samuel O. Oyola, Stephanie N. Seifert. These authors contributed equally (Carolyne Nasimiyu, Damaris Matoke-Muhia, Gilbert K. Rono).

## Abstract

**Background:** Using classical and genomic epidemiology, we tracked the COVID-19 pandemic in Kenya over 23 months to determine the impact of SARS-CoV-2 variants on its progression.

**Methods:** SARS-CoV-2 surveillance and testing data were obtained from the Kenya Ministry of Health, collected daily from 306 health facilities. COVID-19-associated fatality data were also obtained from these health facilities and communities. Whole SARS-CoV-2 genome sequencing were carried out on 1241 specimens.

**Results:** Over the pandemic duration (March 2020 - January 2022) Kenya experienced five waves characterized by attack rates (AR) of between 65.4 and 137.6 per 100,000 persons, and intra-wave case fatality ratios (CFR) averaging 3.5%, two-fold higher than the national average COVID-19 associated CFR. The first two waves that occurred before emergence of global variants of concerns (VoC) had lower AR (65.4 and 118.2 per 100,000). Waves 3, 4, and 5 that occurred during the second year were each dominated by multiple introductions each, of *Alpha* (74.9% genomes), *Delta* (98.7%), and *Omicron* (87.8%) VoCs, respectively. During this phase, government-imposed restrictions failed to alleviate pandemic progression, resulting in higher attack rates spread across the country.

**Conclusions:** The emergence of *Alpha, Delta*, and *Omicron* variants was a turning point that resulted in widespread and higher SARS-CoV-2 infections across the country.

## BACKGROUND

In most countries globally, the COVID-19 pandemic progressed in a series of waves characterized by rapid increase in infection rates followed by a few months of decline before the next wave [1].Factors associated with emergence of new waves included declined application of mitigation measures, climatic changes, and emergence of new virus variants [2,3]. The global genomic surveillance of SARS-CoV-2 played a pivotal role in identifying emerging variants and associated mutations that impacted virus transmissibility, disease severity, vaccine efficacy and clinical case management [3,4]. So far, key variants of public health importance, designated by World Health Organization (WHO) as variants of concern (VoC), had mutations that enhance transmissibility, reduced virus neutralization by antibodies generated following infection or vaccination, interfered with diagnostic testing, and often caused more severe disease [5]. These variants appeared to gain competitive advantage on existing strains to exert immediate global dominance, and most were associated with increased hospitalization or higher mortality, and re-infection of vaccinated or previously infected persons [6].

The paucity of SARS-CoV-2 genomic surveillance data in Africa has limited our understanding of role of virus variants in progression of COVID-19 pandemic in the continent. During the early phase of the pandemic, epidemiologic data from the region suggested lower morbidity and mortality in the region, attributed to factors such as youthful population, favorable weather, and prior exposure to cross-reactive viruses [7]. However, later studies showed infections rates comparable to global trends, but significantly lower levels of severe disease and mortality [8].The emergence of VoCs with global impact in the later phase of the pandemic was associated with increased disease severity, rapid transmission, and re-infection of vaccinated or previously infected persons, continuing to strain the global public health infrastructure and economies despite availability of effective vaccines [9]. Among the VoCs that had global impact were B.1.1.7 (*Alpha*) first identified in United Kingdom in September 2020, B.1.351 (*Beta*) first reported in South Africa in December 2020, B.1.525 (*Eta*) first identified in the UK and Nigeria in December of 2020, B.1.617.1 (*Delta*) first identified in India in October 2020, P.1 (or B.1.1.28.1, *Gamma*) first reported in Brazil in January 2021, and B.1.1.529 (*Omicron*) first reported in South Africa in November 2021[4,10–12] and retroactively detected in samples in the US and other countries around the same time [13]. Two other VOCs, B.1.427 and B1.429 (*Epsilon*) were detected in California, United states in February 2021 but they did not have significant global spread [14].

Progression of COVID-19 pandemic in Kenya may be classified into three phases. The first phase (March 2020-February 2021) started with virus introduction into the country and ended with emergence of VoCs. The second phase (March 2021 – October 2021) was characterized by introduction of various VoCs and vaccination while most COVID-19 restriction remained in place. And the third phase (November 2021 – Present) started when the government lifted most restrictions but also ensured vaccines were widely available. Here, we tracked the pandemic in country over 23 months using classical and genomic surveillance approaches in order to assess the impact of the emerging virus variants on progression of the pandemic. Following confirmation of the first COVID-19 case on March 13, 2020, and through subsequent waves, the government of Kenya implemented various mitigation measures to control its spread, including closure of international borders, banning social gatherings, and lockdown of hotspots located primarily in urban and peri-urban regions of the country [15]). Despite these measures, the SARS-CoV-2 prevalence in capital city of Nairobi was reported as 35% in the first 8 month of the pandemic (in November 2020) [16], and studies predicted that 75% of the Nairobi’s population would be infected by June 2021[17].As at January 30, 2022, Kenya had reported five waves of COVID-19 pandemic, with a total of 331,324 confirmed cases and 5,488 deaths (case fatality ratio = 1.7%)[18]. By then, only 17.3% of the adult population had been vaccinated, in large part due to limited availability of vaccines, and a level of vaccine hesitancy [19–21].

## METHODS

### COVID-19 surveillance data

We abstracted data from the Kenya Ministry of Health (KMOH) COVID-19 daily situation reports between March 12, 2020, and January 30, 2022[18]. The data collected included date of report, number of confirmed cases and deaths at national and county level, age group and sex of cases and deaths, and number of tests conducted. The KMOH situation reports were based on the COVID-19 surveillance system that collected samples from patients presenting at health facilities in 306 sub counties across the entire country and meeting the suspect case definition for COVID-19. The surveillance system also collected samples from healthcare workers with symptoms of a respiratory illness and/or meeting the COVID-19 suspect case definition, people coming in-contact with confirmed COVID-19 cases, and self-initiated testing at 50 biomedical laboratories for a variety of reasons such as heightened suspicion index and international travel.

### SARS-CoV-2 testing and reporting

From each surveilled individual, nasopharyngeal and oropharyngeal swabs were collected, immediately preserved into virus transport medium (VTM), and transported in cool boxes to any of about 200 designated COVID-19 testing laboratories within major hospitals and biomedical research laboratories across the country. In most laboratories, three aliquots of the sample were prepared, and one immediately tested for presence of SARS-CoV-2 virus using RT-PCR. The other two aliquots were transferred to the Sample Management and Receiving Facilities (SMRF) at the Kenya Medical Research Institute (KEMRI) for long-term storage in -80°C. Testing laboratories reported results daily to the National Public Health Laboratories at KMOH through an integrated laboratory information management system.

### SARS-CoV-2 testing inequity

Nationally, the cumulative testing rate was 0.7 tests per 1000 persons per week, against a target of 1 per 1000 persons per week as of January 30, 2022 [18].The testing rate was higher in the major cities of Nairobi, Mombasa, and other urban counties when compared to rural counties. For example, between August and December 2021, the testing rate in Nairobi was 4.7 tests per 1000 persons per week whereas in 9 rural counties the rate was ~0.7 test per 1000 persons per week[18].

## Collection of fatality data

COVID-19 related fatalities occurring within health facilities were reported daily through a standard KMOH death reporting tool developed specifically for the pandemic. Fatalities occurring within the communities were reported directly to 306 Sub-County Disease Surveillance Coordinators (SCDSCs) nationally by the patient’s relatives, community health volunteers, or local government administrators in accordance Kenya Civil Registration Act. All the SCDSCs in the country also collated and reported COVID-19 related mortality data daily to Disease Surveillance and Response Unit at the KMOH national headquarters in Nairobi.

### Epidemiology data analysis

The reported COVID-19 cases and deaths were analyzed by week and county and presented as counts, percentages, ranges, median and in epidemic curves. We defined a wave by epi-week based on three criteria; i) increase in number of reported cases for three consecutive weeks, ii) The start of the wave was the first week of at least three consecutive increases where the increase from the previous week was at least 35%, and iii) the end of the wave was defined as the week when the reported cases were equal to or lower than those reported during the week of onset of the wave. The attack rate was defined as the number of reported cases divided by the human population at national and county level. Case fatality ratio (CFR) was defined as the ratio of deaths to reported cases. We estimated the 95% confidence interval of the proportion of cases and deaths on the binomial distribution defined by the observed proportions.

### SARS-CoV-2 genomic surveillance

Starting from May 2020, we selected up to 200 real-time PCR-positive specimens with CT ≤32.0 per month from the KEMRI SMRF for whole genome sequencing of SARS-CoV-2 at either the Regional Genomic Center of International Livestock Research Institute (ILRI) or Center for Biotechnology Research and Development of KEMRI.

### Whole genome sequencing and variant identification

The SARS-CoV-2 whole genome sequencing was carried out as described previously [22].Briefly, viral RNA was extracted from sample either manually or using TANBead® Maelstrom 9600 (Taiwan Advanced Nanotech Inc, Taiwan) automated nucleic acid extractor according to the manufacturer’s directions. For manual extraction, 140 ul of sample was applied in the QIAGEN QIAamp® Viral RNA Mini Kit (Hilden Düsseldorf, Germany). RNA was eluted in 60µl buffer and stored in RNAse-free Eppendorf tubes. The NEBNext-Artic SARS-CoV-2 library preparation workflows for both Illumina and Oxford Nanopore Technologies (ONT) were used[23]. For Illumina, the protocol NEBNext® ARTIC SARS-CoV-2 Library Prep Kit (Illumina®) (Version 2.0_3/21) was used following manufacturer’s instructions.

Sequencing was done on the Illumina MiSeq or NextSeq 550 platforms. Demultiplexing and adapter trimming were performed automatically by the sequencing onboard software. For ONT, the ARTIC SARS-CoV-2 Library Prep Kit (ONT®) were used. There was minimal deviation with both Illumina and ONT workflows. Size distribution was estimated using agarose gel electrophoresis. ONT sequencing was done on the MinION platform. Base calling, demultiplexing and adapter trimming was performed using Guppy v5.0.11 and fastq outputs used for downstream analyses.

Variant calling and lineage/clade assignment were carried out using the singularity container of the nf-core/viralrecon v2.2: an analysis pipeline for assembly and intra-host/low-frequency variant calling for viral samples[24]. Majorly, for Illumina, we used Fastp to trim low-quality reads; Bowtie for read mapping, and iVar for removal of primer sequences, mutation calling and for consensus generation. While with ONT data, read mapping and consensus generation was by minimap2, and medaka was employed for mutation calling. In both cases, snpEff was used for mutation annotation. The nf-core/viral recon ONT workflow embeds the ARTIC ONT pipeline. Further downstream, consensus sequences were used by Pangolin USHER for lineage assignments based on parsimony and Nextclade [25] for clade specification.

### Phylogenetic analyses

Apart from our sequences (n=1241), we downloaded an additional 894 complete SARS-CoV-2 genomes from various lineages for phylogenomic comparison. To compare with global sequences, consensus sequences were aligned using NextAlign, embedded in Nextclade, and resulting multiple sequence alignment (MSA) fed into IQ-TREE [26] for inferring maximum likelihood to determine the most likely phylogram describing the combined dataset. Tree visualization and annotation were done using the FigTree software [27]. For time-resolved phylogenetic trees, we used Mafft aligner v1.10.0[28] to generate MSA of the 1241 genomes. To account for homoplasy that may affect our phylogeny, we used the ClonalFrameML [29] to estimate the phylogeny with corrected branch lengths.

Lineage designation was implemented in pangolin v 3.0 .1.17 [30] with USHER mode for parsimony-based lineage assignment [31]. Multiple sequence alignment was performed on SARS-CoV-2 sequences from all samples while separate alignments were performed for *Delta* variants using NextAlign[32]. For both alignments, the maximum likelihood phylogenetic tree was inferred using IQ-TREE v 2.1.3 [26] with ModelFinder [28] and 1000 UltraFast bootstrap replicate approximation[33]. The time-resolved phylogenetic tree for all Kenya sequences was then inferred in TreeTime [34] with the Wuhan-Hu-1 variant (NCBI reference: NC_045512.2) as the outlier. Phylogenetic trees were visualized with package ggtree [35] implemented in R v 4.1.2. The new SARS-CoV genomes sequenced in this study were submitted to either global initiative on sharing avian influenza data (GISAID, https://www.gisaid.org/) or National Center for Biotechnology Information (NCBI, https://www.ncbi.nlm.nih.gov/) and accession numbers provided in **Supplement Table 1** and **Supplement Table 2**, respectively.

### Ethical approvals

The COVID-19 surveillance and testing data was collected from public database of the Kenya Ministry of Health (KMOH) with administrative approval from the ministry. Genomic surveillance study was reviewed and approved by the KEMRI Scientific and Ethical Review Unit (KEMRI/SERU/CVR/006/4035), ILRI Institutional Research Ethics Committee (ILRI-IREC2020-52), and a reliance approval provided by Washington State University Institutional Review Board based on in-country ethical reviews as provided for in Code of Federal Regulations (45 C.F.R part 46 and 21 C.F.R. part 56). Data analysis and manuscript development were in partnership with KMOH and KEMRI as demonstrated by authorship.

## RESULTS

### Pandemic waves and regional spread of infections

Between March 2020 and January 2022, Kenya experienced five waves of COVID-19 (**Figure 1**). Wave-1 and wave-2 occurred during the first phase of the pandemic before global emergence of VoCs, wave-3 and wave-4 during second phase when VoCs emerged and vaccination started, and wave-5 during the current phase of the pandemic after Kenya had lifted most restrictions. Wave-2 was the longest at 10 weeks, while wave-1, wave-3, and wave-4 lasted for 8 weeks each, and wave-5 for 7 weeks. The shortest inter-wave period of 4 weeks was observed between waves 1 and 2, while those between waves 2 and 3, waves 3 and 4, and waves 4 and 5 were 8 to 11 weeks long (**Figure 1**). Reported cases between wave-3 and wave-4 were higher when compared to the cases reported between the other waves.

**Figure 1.**
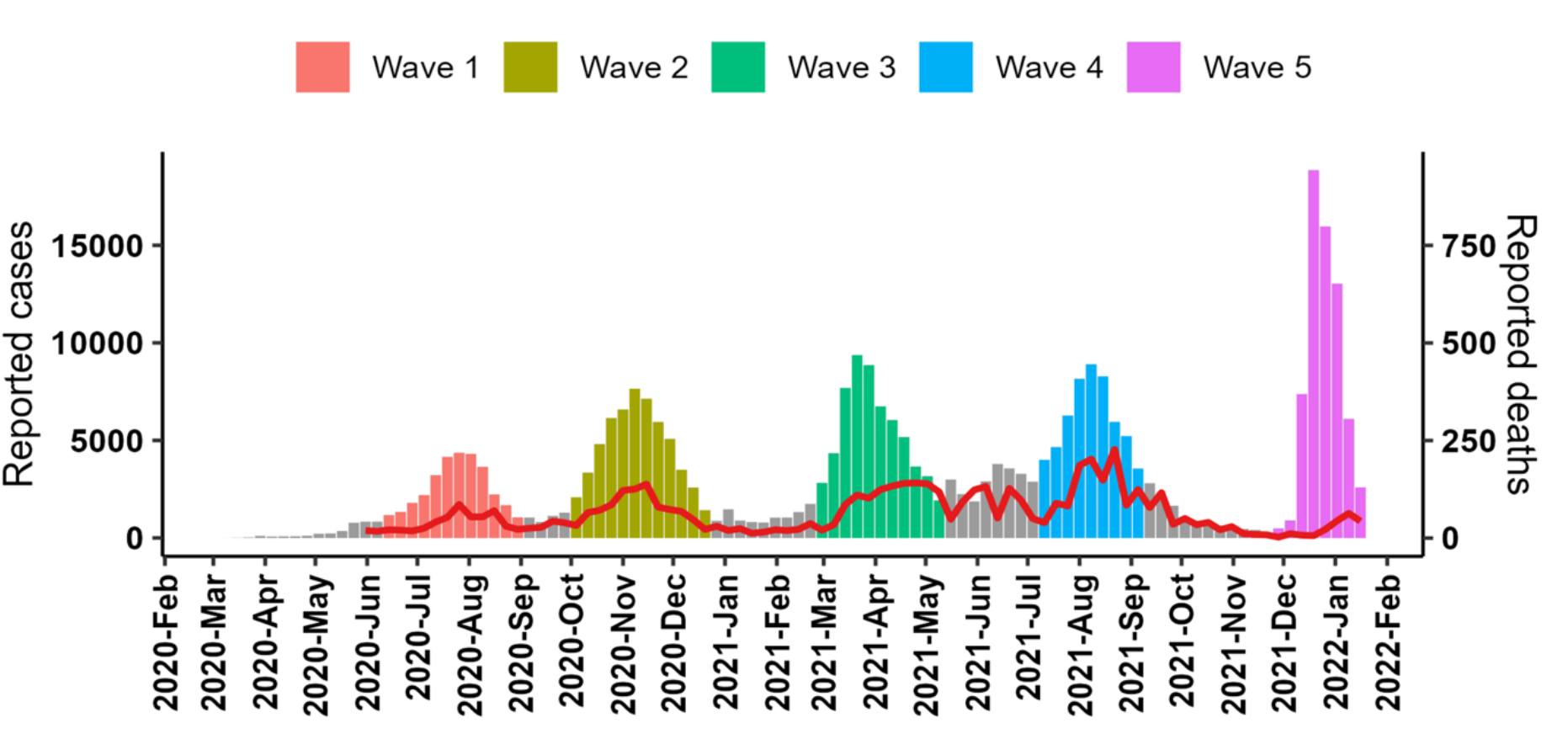
Number of reported COVID-19 cases and deaths by epidemiological week, 2020-2022, Kenya. The five waves are highlighted in different colours. The numbers of fatalities are denoted by the red line graph with a secondary axis to the right.

The national attack rate (AR) during the waves ranged from 65.4 to 137.6 cases per 100,000 persons with the highest AR reported in wave-5 and the lowest in wave-1. During wave-1, the median AR per county was 14.6 (Inter-quartile range =29.9) cases per 100,000 persons across the countries 47 counties, with only 4 counties surrounding Nairobi city in southcentral Kenya, and Mombasa city along the Indian Ocean coastal region reporting AR >100 cases per 100,000 persons (**Figure 2**).In contrast, during wave-3 and wave-4, the median AR rate was 70.3 (IQR = 98.5) cases per 100,000 persons, with 17 (36.2%) of the counties reporting AR >100 cases per 100,000 persons. Overall, Nairobi city accounted for 43% of all the cases reported during the five waves (range 36.9% -60.7%), and the highest intra-wave AR ranging between 460.9 and 627.2 per 100,000 persons.

**Figure 2.**
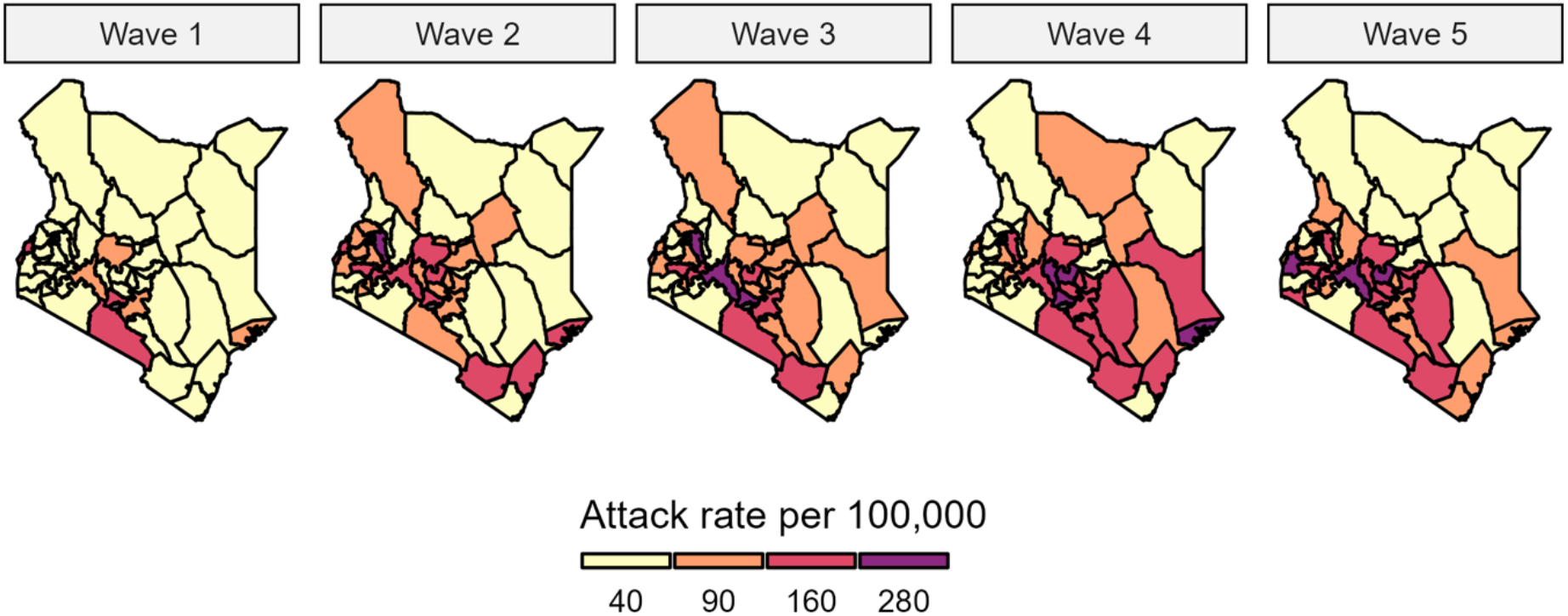
SARS-CoV-2 attack rate by county and wave in Kenya.

### Case fatality ratio

The number of COVID-associated deaths reported was higher during the waves when compared to the number reported between the waves (**Figure 1**). Whereas the average CFR over the 23-month pandemic period was 1.7%, the intra-wave CFR 3.5%. Interestingly, while the intra-wave CFR was between 3.9% and 6.6% during waves 1-4, it dropped to 0.3% in wave 5. People aged below 20 years, who constitute >50% of the population contributed 10.0% of cases but only 2.9% of deaths. In contrast, people aged ≥60 years old (4.2% of the population) contributed 13.7% of the cases and 56.5% of deaths (**Table 1**). Although only 26.6% of the cases occurred in persons >49 years, this age group contributed 74.3% of the deaths. Reported deaths among males were almost twice higher than those reported among females (**Table 1**).

**Table 1:** Categorization of COVID-19 cases and deaths by age groups and sex in Kenya

|  | All Cases |  | All deaths |  | Female cases | Male cases | Male deaths | Female deaths |
| --- | --- | --- | --- | --- | --- | --- | --- | --- |
| Age group | n | % (95% CI) | n | % (95% CI) | % (95% CI) | % (95% CI) | % (95% CI) | % (95% CI) |
| 0-9 | 12064 | 3.8(3.7-3.8) | 89 | 1.6( 1.3-2) | 43.7(42.8-44.6) | 56.3(55.4-57.2) | 65.2(54.3-74.8) | 34.8(25.2-45.7) |
| 10-19 | 19832 | 6.2(6.1- 6.3) | 72 | 1.3(1-1.6) | 47.6(46.9-48.3) | 52.4(51.7-53.1) | 58.3(46.1-69.6) | 41.7(30.4-53.9) |
| 20-29 | 60860 | 19(18.9-19.1) | 162 | 2.9(2.5-3.4) | 47.3(46.9-47.7) | 52.7(52.3-53.1) | 51.9(43.9-59.7) | 48.1(40.3-56.1) |
| 30-39 | 83067 | 25.9(25.8-26.1) | 428 | 7.7(7-8.4) | 43.1(42.8-43.5) | 56.9(56.5-57.2) | 51.9(47-56.7) | 48.1(43.3-53) |
| 40-49 | 59099 | 18.5(18.3-18.6) | 683 | 12.2(11.4-13.1) | 40.1(39.7-40.5) | 59.9(59.5-60.3) | 63.5(59.8-67.1) | 36.5(32.9-40.2) |
| 50-59 | 41394 | 12.9(12.8-13) | 996 | 17.8(16.9-18.9) | 41.5(41.1-42) | 58.5(58-58.9) | 68.4(65.4-71.2) | 31.6(28.8-34.6) |
| ≥60 | 43998 | 13.7(13.6-13.9) | 3152 | 56.5(55.2-57.8) | 44.2(43.8-44.7) | 55.8(55.3-56.2) | 64.6(62.9-66.3) | 35.4(33.7-37.1) |
| Total | 320314 | - | 5582 | - | 43.6(43.4-43.8) | 56.4(56.2-56.6) | 63.7(62.5-65) | 36.3(35-37.5) |

### Dominant SARS-CoV-2 lineages during waves

The 1241 SARS-CoV-2 genomes sequenced between May 2020 and January 2022 were assigned to 24 distinct Pango lineages with the most common lineages being B.1.617.2 (*Delta*, 38.4%), B.1(non-VOC, 24.6%), B.1.1.7 (*Alpha*, 16.5%), and B.1.1.529 (*Omicron*, 7.5%).

During the first phase of pandemic, the B.1 global parental lineage, which circulated from the beginning of the pandemic in the country, dominated accounting for 94% of all genomes in wave-1, and 71% in wave-2 (**Figure 3**). Diverse virus variants started emerging in wave-2 through to wave-3, both VoCs such as B.1.351 (*Beta*) and B.1.1.7 (*Alpha*) and non-VoCs such as B.1.3x, B.1.5x, B.1.525 (*Eta*), A and A.23x. However, midway through wave-3, *Alpha* emerged as dominant variant accounting for 74.9% of all genomes sequenced (**Figure 3 & 4**). The B.1.617.2 (*Delta*) and its sub-variant AY.x were first detected in March 2021 and swept away other variants to become the dominant variant (99.3% of the genomes sequenced) during wave-4 (**Figures 3 & 4**).

**Figure 3.**
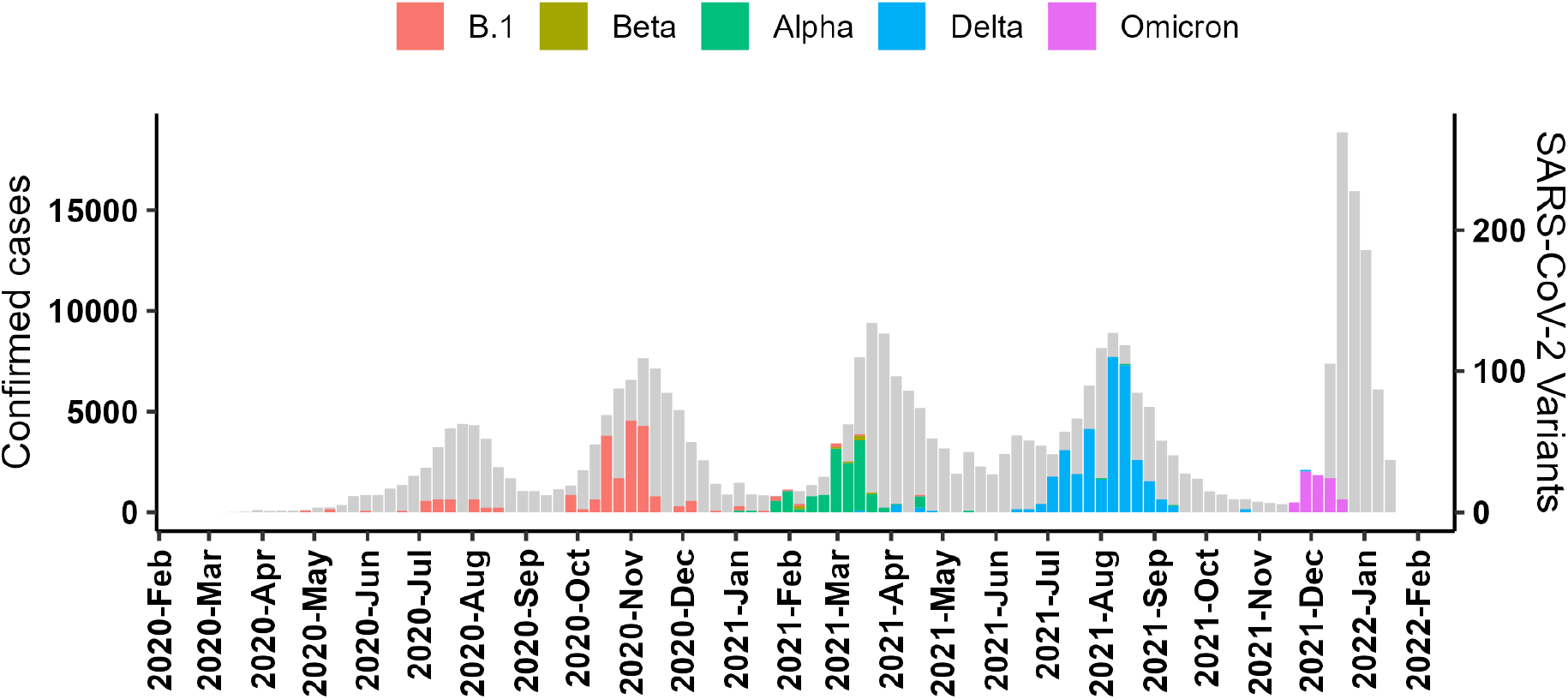
Kenyan COVID-19 Epi-curve as of January 30, 2022, showing the dominant SARS-CoV-2 variants during each of the 5 waves.

**Figure 4.**
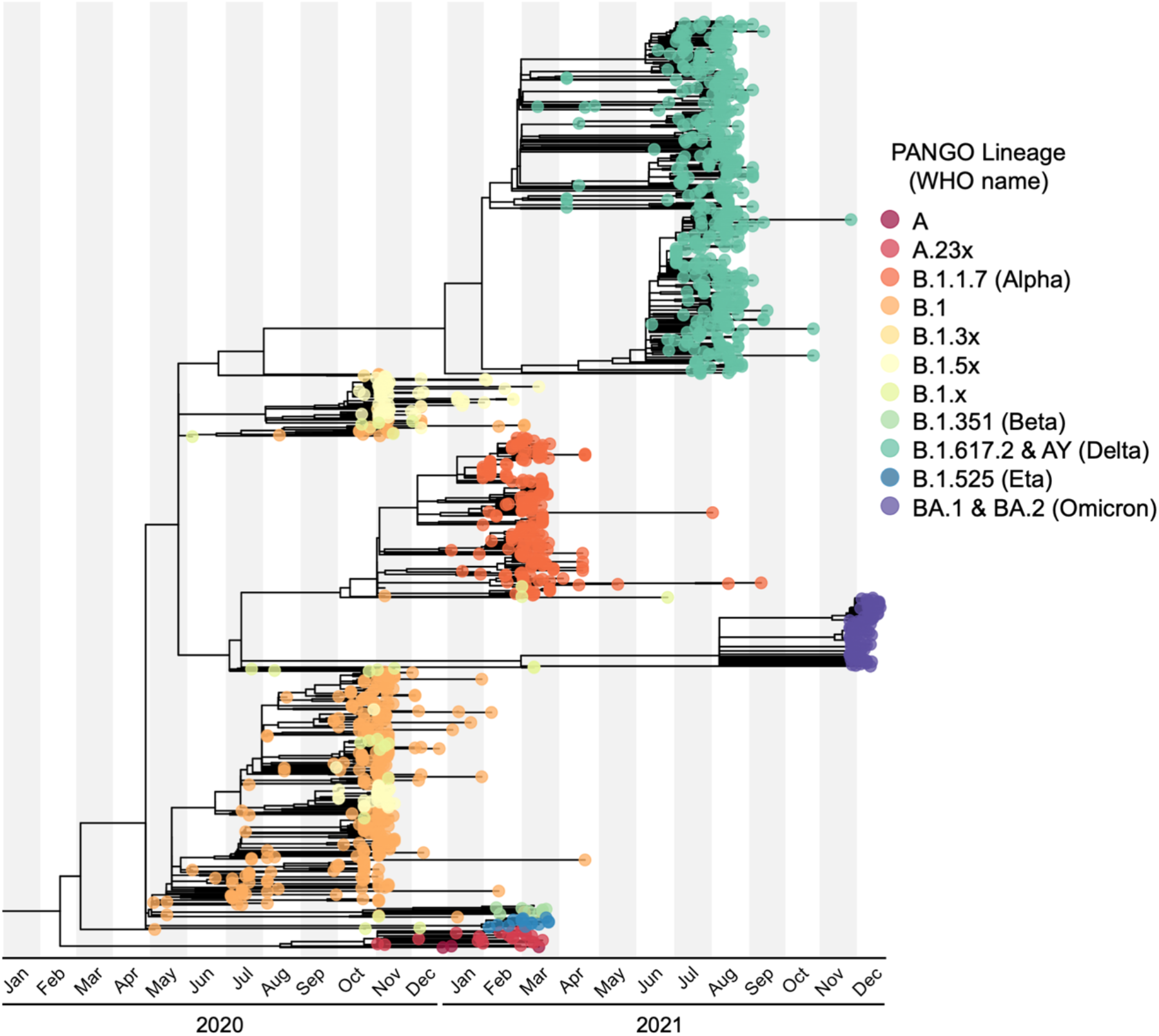
A time calibrated phylogenetic tree of 1241 SARS CoV-2 genomes sequenced from Kenyan patients between May 2020 and December 2022. Pango lineage designations are indicated by the circles on the branch tips, including VoCs and variants of interest as designated by World Health Organization. Using the 2019 Wuhan-Hu-1 genome (GenBank accession number MN908947.3) as the root of the tree.

The B.1.1.529 (Omicron) lineage was first detected in Kenya in November 20, 2021 and by mid-December it’s sub-variant BA.1 become dominant accounting for 87.8% of all genomes sequenced (**Figures 3 & 4**). Of the major VoCs, only *Beta, Alpha, Delta*, and *Omicron* were detected in the Kenya samples sequenced. Genetic evolution analysis showed intra-lineage diversity of various variants (**Figure 5**). Of the VoCs, *Delta* variant showed greater genetic diversity, including multiple globally circulating AY.X lineages, consistent with multiple introductions as depicted by the three divergent clusters (**Supplemental Figure 1, Figures 4 & 5**). This analysis indicates that the *Alpha* variant and the *Omicron* have a closer common ancestor compared to the *Delta* variant which appears to have diverged from a more distant ancestor (**Figure 5**).

**Figure 5:**
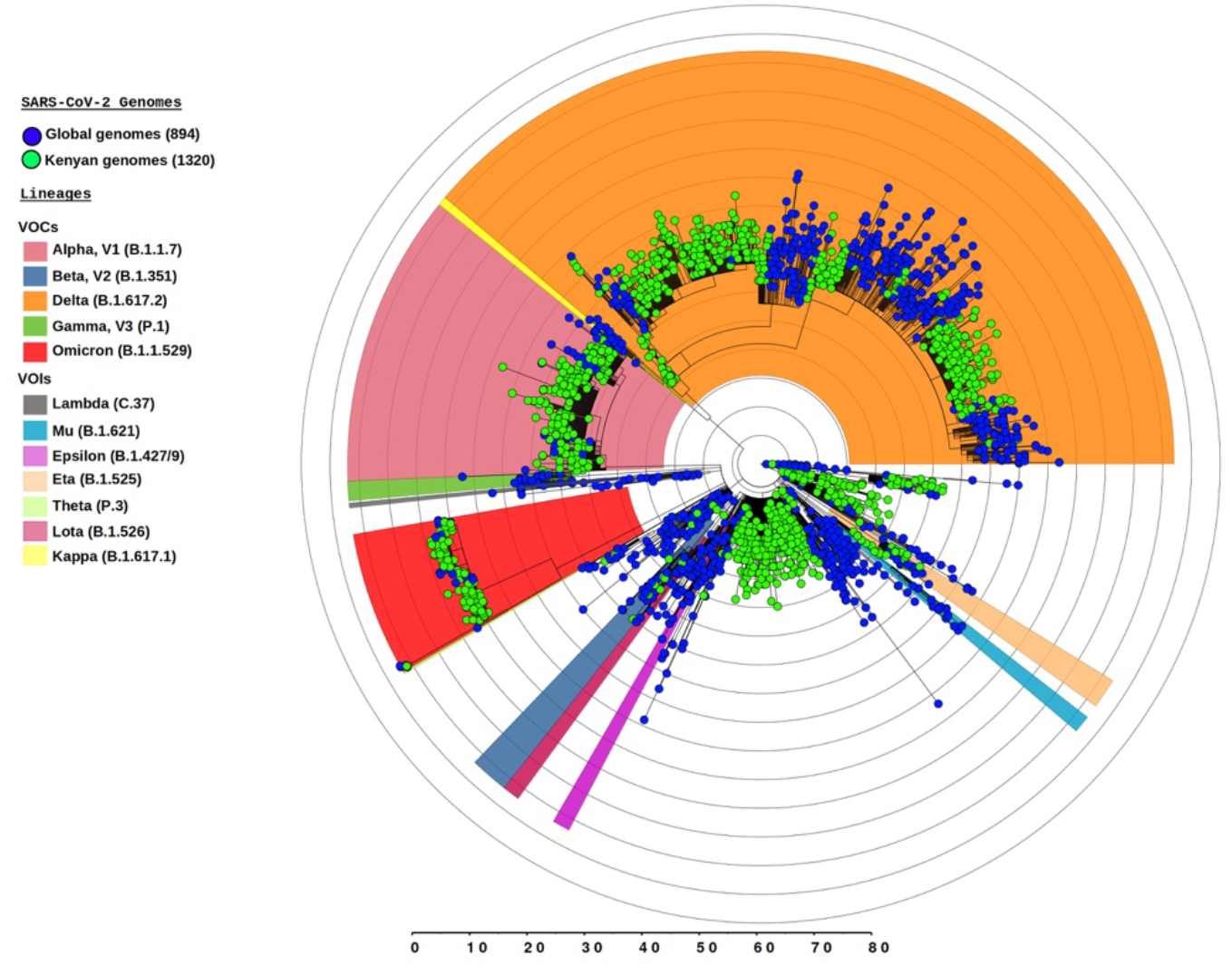
Circular phylogenetic tree depicting evolutionary relationship of Kenyan SARS-CoV-2 sequences (n=1241) against global genomes(n=894) clustered by pangolin lineage annotation as shown in different background colours. The tree is rooted with the Wuhan (MN908947.3) reference genome. Kenyan genomes are shown in green circular tips while the global genomes are shown in blue tips. Lineages representing the major global variants of concern (VoC) detected in Kenya are shown: B.1.1.7 (*Alpha*) is shown in pick background; B.1.351(Beta) is shown in deep blue background; (B.1.617.2 (*Delta*) is shown in orange background; B.1.1.529 (*Omicron*) is shown in red background.

## DISCUSSION

We used both classical and genomic epidemiologic approaches to track the COVID-19 pandemic in Kenya over 23 months (March 2020-January 2022) and assess the impact of emerging virus variants on pandemic progression and severity in the country. In the first phase of the pandemic, the country experienced two waves, characterized by national AR of between 65.4 and 118.2 per 100,000 persons (Nairobi 404 – 474 cases per 100,000 persons), and CFR of 3.9 – 4.2%. The B.1 lineages of the virus dominated, except toward the end of that period (January 2021) when *Alpha*, the first VoC in the country, was detected. During this period, most cases were reported within and around the two main ports of entry into Kenya, the capital city of Nairobi in the southcentral region that received most of the international traveller, and Mombasa along the Indian ocean where most cargo deliveries to the east Africa region are received (Figure 2).

The government responded to the waves by implementing various mitigation measures including closure of borders, in-person schooling closures, and ban on social gatherings. Most of these measures remained in place through October 2021 (18 months into the pandemic), but they became less effective in preventing widespread infections in the country in subsequent months when global VoCs started to emerge. The second phase (March – October 2021) was characterized by emergence of imported VoCs and introduction of COVID-19 vaccines in the country. Early in this period, *Alpha* and shortly after *Delta* variants emerged and were associated with two major waves (wave-3 & wave-4). The *Delta* variant dominated through 5 of the 8 months in this period, and the two waves were associated with the high AR (national 115.6 – 125.7 per 100,000 persons, Nairobi 457.5 – 614.2 per 100,000 persons) and CFR (up to 6.6%). This period saw spread of infections across all 47 counties in the country, but with the higher AR reported in Nairobi and its surrounding counties. In addition to the early mitigation measures, lockdowns were introduced for over 2 months in defined hotspot of the country characterized by high AR. Interestingly, the March-October 2021 period was also marked by introduction of COVID-19 vaccines, albeit slowly, because of low vaccine availability in low-income countries globally. During that 8 month-period, only 19% of the 27 million eligible Kenyans had received at least one dose of the vaccine [18].

Since October 2021, Kenya has been in the third phase of pandemic characterized by lifting of most restriction to re-open the economy and increased availability of vaccines. During this phase in December 2021, the *Omicron* variant emerged, creating wave-5 associated with the highest AR (national 137.6 per 100,000 persons, Nairobi 627.3 per 100,000 persons) but lowest CFR (CFR = 0.3%). The government did not re-introduce restrictions, and vaccines became more widely available. By the end of January 2022, 42% of eligible Kenyans had received at least one dose of the vaccine, but only 0.4% had received the recommended 3^rd^ booster doses [36]. Overall, more deaths were experienced among the elderly people (60 years and above) compared to the younger people, a result comparable to global trends.

Of the >2,800 SARS-CoV-2 genomes reported from Kenya in the current and other recent studies [37,38], there has been no VoC emerging from the country, with only B.1.525 (*Eta*) that was detected in February 2021 in the Nairobi classified as variant of interest. Studies point at increased transmissibility and capacity to evade the immune response as the key factors associated with dominance of the VoCs [39,40]. There is raging public debate associating vaccine inequity with emergence of these variants; however, the evidence so far remains inconclusive. For instance, emergence of Delta from India and Omicron from South Africa, countries that had low vaccination coverage at the time, appears to support this hypothesis. However, the fact that we have not seen many VoCs emerging from Kenya and other African countries apart from South Africa, most of them with <20% vaccine coverage by end of 2021 does not support the argument. Studies suggest that new VoCs can competitively gain advantage over existing variants through various mechanisms, including having higher infectivity, longer duration of infection, or being less virulent to cause asymptomatic disease that is harder to detect [6,41,42]. Breakthrough infections of vaccinated individuals have been widespread with *Omicron*, however, a complete vaccination regimen that includes the third booster dose improves virus neutralization against the *Omicron* variant and may result in reduced transmission[43].

Our genomic analyses suggest multiple introductions of imported SARS-CoV-2 variants from both regional and international sources. Though later waves were dominated by a single imported VoC (*Alpha, Delta*, or *Omicron*), we detected subtypes of these major variants associated with the US, Europe, India, Nigeria, DRC, and Uganda, supporting multiple introductions. There are nearly 8 million SARS-CoV-2 genomes available in GISAID and more than 50% of these sequences originate from just two regions, the North America and Europe. Only about 1% of SARS-CoV-2 genomes are from Africa. Of these Africa genomes, nearly 40% of sequences originate from South Africa (GISAID, accessed Feb 2^nd^, 2022). This sampling bias reduces the likelihood that most African countries would have detected emerging variants with consequential mutations in a timely manner. However, we expect that if a VoC with sustained global impact emerged in the region, it could have been detected through clinical disease profile or outside Africa in countries with more robust genomic surveillance. The inter-wave duration ranged between 8-11 weeks for wave-2 through to wave-5, perhaps suggesting time to allow population immunity to wane and support emergence of the next wave. Interestingly, the inter-wave period between wave-1 and wave-2 was only 4 weeks, likely reflecting the low level of the population immunity to SARS-CoV-2 at the time.

A limitation of the study is that reported COVID-19 cases and deaths and samples used for genomic surveillance were based on the KMOH surveillance, and which likely underestimated the extent of the pandemic at any one time. Nonetheless, surveillance plays a critical role in monitoring trends and control of global pandemics.

In conclusion, the emergence of *Alpha, Delta*, and *Omicron* VoCs was a turning point that resulted in widespread and higher SARS-CoV-2 infections across the country, with varying fatality rates. Enhanced genomic and molecular surveillance for SARS-CoV-2 in developing countries can support early detection of VoCs and guide public health control measures such application of mitigation measures.

## Data Availability

All data produced are available online at GISAID or NCBI or through the Kenya Ministry of Health.

## ACKNOWLEDGEMENTS

We thank Shebbar Osiany, Paul Dobi, Edward Nyaga, Collins. Muli, Regina Njeru, Reuben Mwangi for supporting genomic work, Missiani Ochwoto, Ruth Cheruto, James Kimotho, and Jeremiah Omar for supporting testing, and the Washington State University Global Health Kenya staff for administrative support. The findings and conclusions in this study are those of the authors and do not necessarily represent the official position of the Kenya MOH, KEMRI, US NIH, or CGIAR/ILRI.

---

**Supplement Figure 1.**
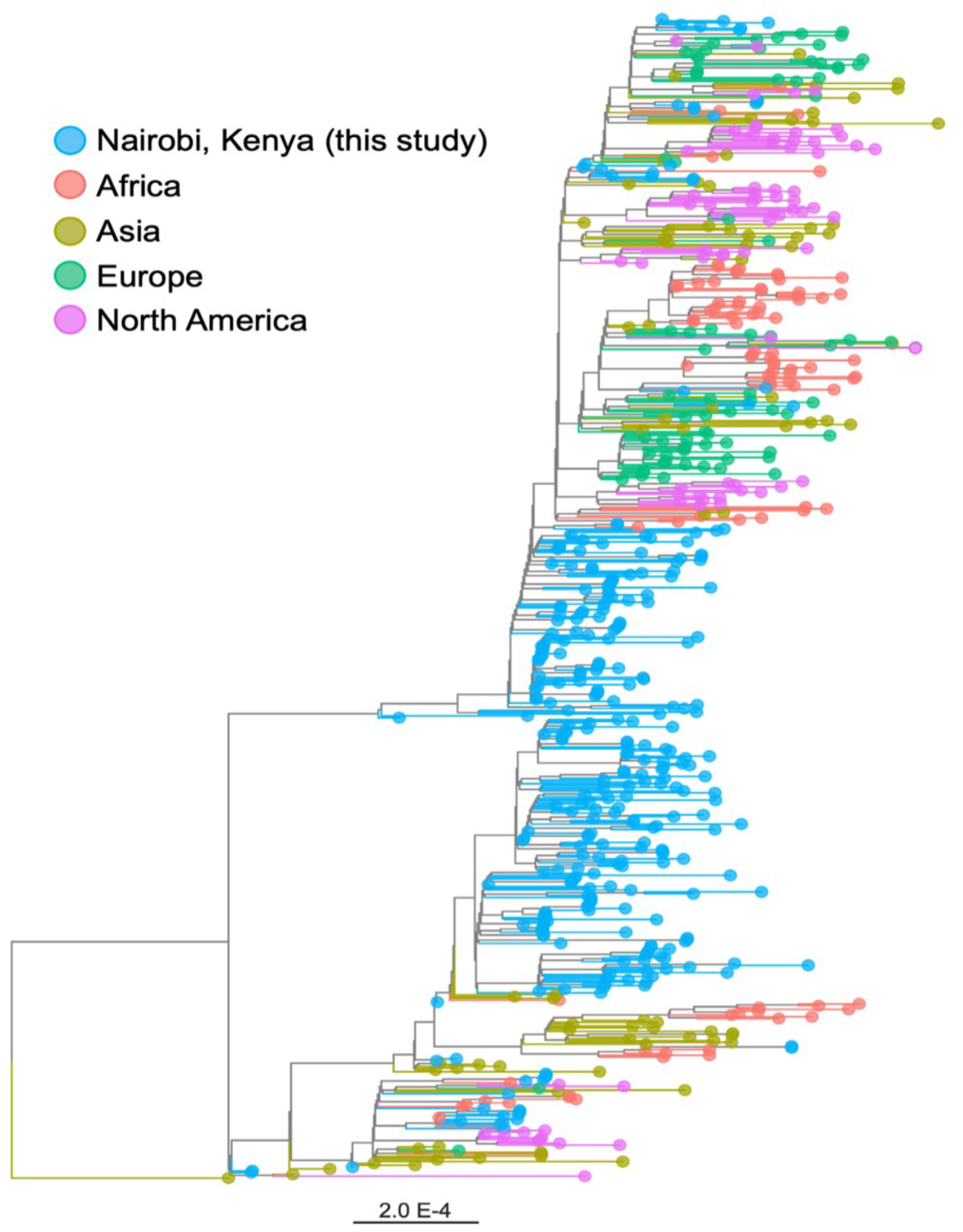
Maximum likelihood tree showing global context of SARS-CoV-2 Delta variants circulating in Nairobi, Kenya. Region of origin for each SARS-CoV-2 variant is indicated by the colour of the circles at the branch tips.

**Supplement Table 1**. Accession number for the 1179 genomes sequenced in this study submitted to global initiative on sharing avian influenza (GISAID) database (https://www.gisaid.org/).

**Supplement Table 2**. Accession number for the 62 genomes sequenced in this study submitted to National Center for Biotechnology Information (NCBI) database (https://www.ncbi.nlm.nih.gov/).

