## Supplemental Tables 1 and 2 for "Imported SARS-COV-2 Variants of Concern Drove Spread of Infections Across Kenya During the Second Year of the Pandemic": SupplementalTables.pdf

| **Supplement Table 1.** Accession Numbers for SARS-CoV genomes submitted to GSAID | | | | | |
|---|---|---|---|---|---|
| **Local Name** | **Accession Number** | **Local Name** | **Accession Number** | **Local Name** | **Accession Number** |
| COVM01389 | EPI_ISL_6096565 | COVC19665 | EPI_ISL_4252311 | COVC22974 | EPI_ISL_8318551 |
| COVM01392 | EPI_ISL_6096566 | KEM-21-03-93912 | EPI_ISL_4252312 | COVC16122 | EPI_ISL_8318557 |
| COVM01384 | EPI_ISL_6096541 | COVC20832 | EPI_ISL_4252313 | KEM-21-03-95378 | EPI_ISL_8318556 |
| COVM01393 | EPI_ISL_6096574 | COVC20841 | EPI_ISL_4252316 | COVC23644 | EPI_ISL_8318562 |
| COVM01394 | EPI_ISL_6096579 | COVC04131 | EPI_ISL_4252317 | COVC21480 | EPI_ISL_8318559 |
| COVM01397 | EPI_ISL_6096580 | COVC03965 | EPI_ISL_4252319 | COVC23001 | EPI_ISL_8318568 |
| COVM00453 | EPI_ISL_6093674 | COVC13005 | EPI_ISL_4252323 | COVC14960 | EPI_ISL_8318565 |
| COVM00457 | EPI_ISL_6093680 | COVC13752 | EPI_ISL_4252326 | KEM-21-03-95374 | EPI_ISL_8318567 |
| COVM00468 | EPI_ISL_6093709 | COVC21366 | EPI_ISL_4252327 | COVC22979 | EPI_ISL_8318569 |
| COVM00471 | EPI_ISL_6093713 | COVC04554 | EPI_ISL_4252329 | COVC23696 | EPI_ISL_8318539 |
| COVM00472 | EPI_ISL_6093722 | COVC19900 | EPI_ISL_4252330 | COVC23319 | EPI_ISL_8318535 |
| COVM00474 | EPI_ISL_6093730 | COVC23946 | EPI_ISL_4252332 | COVC23286 | EPI_ISL_8318542 |
| COVM00475 | EPI_ISL_6093736 | COVC23960 | EPI_ISL_4252334 | COVC14815 | EPI_ISL_8318545 |
| COVM00476 | EPI_ISL_6093738 | KEM-21-03-94392 | EPI_ISL_4252335 | COVC16127 | EPI_ISL_8318543 |
| COVM00498 | EPI_ISL_6093759 | COVC23888 | EPI_ISL_4252336 | COVC23346 | EPI_ISL_8318549 |
| COVM00501 | EPI_ISL_6093761 | KEM-21-03-93952 | EPI_ISL_4252338 | COVC22895 | EPI_ISL_8318528 |
| COVM00502 | EPI_ISL_6093762 | KEM-21-03-95523 | EPI_ISL_4252339 | COVC24016 | EPI_ISL_8318610 |
| COVM00504 | EPI_ISL_6093775 | COVC16084 | EPI_ISL_4252346 | COVC23693 | EPI_ISL_8318617 |
| COVM00509 | EPI_ISL_6093776 | COVC22803 | EPI_ISL_4252350 | COVC16215 | EPI_ISL_8318612 |
| COVM00516 | EPI_ISL_6093783 | COVC24020 | EPI_ISL_4252351 | COVC23551 | EPI_ISL_8318609 |
| COVM00533 | EPI_ISL_6093798 | KEM-21-03-95532 | EPI_ISL_4252353 | COVC23609 | EPI_ISL_8318614 |
| COVM00535 | EPI_ISL_6093799 | COVC24018 | EPI_ISL_4252354 | COVC24017 | EPI_ISL_8318601 |
| COVM00538 | EPI_ISL_6093805 | COVC24170 | EPI_ISL_4252355 | KEM-21-03-94652 | EPI_ISL_8318520 |
| COVM00539 | EPI_ISL_6093812 | KEM-21-03-95582 | EPI_ISL_4252356 | COVC23671 | EPI_ISL_8318607 |

| | | | | | |
|---|---|---|---|---|---|
| COVM00540 | EPI_ISL_6093813 | KEM-21-03-93872 | EPI_ISL_4252359 | KEM-21-03-94501 | EPI_ISL_8318516 |
| COVM00541 | EPI_ISL_6093815 | COVC13799 | EPI_ISL_4252361 | KEM-21-03-94391 | EPI_ISL_8318519 |
| COVM00543 | EPI_ISL_6093818 | COVC13798 | EPI_ISL_4252366 | COVC16141 | EPI_ISL_8318524 |
| COVM00544 | EPI_ISL_6093825 | COVC03665 | EPI_ISL_4252370 | COVC23617 | EPI_ISL_8318523 |
| COVM00546 | EPI_ISL_6093826 | COVC20838 | EPI_ISL_4252377 | COVC15802 | EPI_ISL_8318525 |
| COVM00548 | EPI_ISL_6093836 | COVC24155 | EPI_ISL_4252378 | COVC20037 | EPI_ISL_4252048 |
| COVM00552 | EPI_ISL_6093843 | COVC23356 | EPI_ISL_4252380 | COVC03723 | EPI_ISL_4251929 |
| COVM00553 | EPI_ISL_6093845 | COVC23562 | EPI_ISL_4252381 | COVC03955 | EPI_ISL_4251930 |
| COVM00556 | EPI_ISL_6093853 | COVC21313 | EPI_ISL_4252383 | COVC03960 | EPI_ISL_4251931 |
| COVM00558 | EPI_ISL_6093862 | COVC20817 | EPI_ISL_4252391 | COVC04111 | EPI_ISL_4251932 |
| COVM00561 | EPI_ISL_6093870 | COVC23969 | EPI_ISL_4252393 | COVC24001 | EPI_ISL_4252015 |
| COVM00564 | EPI_ISL_6093882 | COVC20846 | EPI_ISL_4252397 | COVC04555 | EPI_ISL_4251935 |
| COVM00567 | EPI_ISL_6093889 | COVC23367 | EPI_ISL_4252400 | COVC13751 | EPI_ISL_4251946 |
| COVM00568 | EPI_ISL_6093895 | COVC23201 | EPI_ISL_4252401 | COVC14832 | EPI_ISL_4251947 |
| COVM00570 | EPI_ISL_6093903 | COVC18976 | EPI_ISL_4252403 | COVC21059 | EPI_ISL_4251951 |
| COVM00571 | EPI_ISL_6093910 | COVC23314 | EPI_ISL_4252412 | COVC21310 | EPI_ISL_4251955 |
| COVM00574 | EPI_ISL_6093919 | COVC18965 | EPI_ISL_4252414 | COVC21039 | EPI_ISL_4251985 |
| COVM00576 | EPI_ISL_6093928 | COVC15018 | EPI_ISL_4252424 | COVC22784 | EPI_ISL_4251986 |
| COVM00577 | EPI_ISL_6093935 | COVC16123 | EPI_ISL_4252428 | COVC23432 | EPI_ISL_4251987 |
| COVM00579 | EPI_ISL_6093937 | COVC16147 | EPI_ISL_4252429 | COVC23192 | EPI_ISL_4251993 |
| COVM00581 | EPI_ISL_6093951 | COVC18954 | EPI_ISL_4252432 | NHRL-S002 | EPI_ISL_4251996 |
| COVM00582 | EPI_ISL_6093956 | COVC23742 | EPI_ISL_4252433 | COVC19906 | EPI_ISL_4252000 |
| COVM00584 | EPI_ISL_6093964 | COVC21028 | EPI_ISL_4252436 | COVC19976 | EPI_ISL_4252002 |
| COVM00589 | EPI_ISL_6093975 | COVC04223 | EPI_ISL_4252446 | COVC21367 | EPI_ISL_4252004 |
| COVM00590 | EPI_ISL_6093980 | COVC19909 | EPI_ISL_4252451 | KEM-21-03-94006 | EPI_ISL_4252010 |
| COVM00591 | EPI_ISL_6093981 | COVC07067 | EPI_ISL_4252452 | COVC22821 | EPI_ISL_4252013 |
| COVM00600 | EPI_ISL_6093997 | COVC21332 | EPI_ISL_4252453 | COVC20863 | EPI_ISL_4252014 |

| | | | | | |
|---|---|---|---|---|---|
| COVM00603 | EPI_ISL_6094003 | COVC19039 | EPI_ISL_4252458 | COVC14802 | EPI_ISL_4252021 |
| COVM00610 | EPI_ISL_6094018 | COVC19983 | EPI_ISL_4252461 | COVC23182 | EPI_ISL_4252023 |
| COVM00611 | EPI_ISL_6094025 | COVC23513 | EPI_ISL_4252465 | COVC15801 | EPI_ISL_4252024 |
| COVM00615 | EPI_ISL_6094035 | COVC21031 | EPI_ISL_4252470 | COVC20036 | EPI_ISL_4252025 |
| COVM00645 | EPI_ISL_6094076 | COVC18974 | EPI_ISL_4252473 | COVC16129 | EPI_ISL_4252026 |
| COVM00649 | EPI_ISL_6094083 | KEM-21-03-95380 | EPI_ISL_4252474 | COVC15079 | EPI_ISL_4252027 |
| COVM00651 | EPI_ISL_6094090 | KEM-21-03-94645 | EPI_ISL_4252475 | COVC19815 | EPI_ISL_4252036 |
| COVM00652 | EPI_ISL_6094096 | COVC18948 | EPI_ISL_4252477 | COVC21224 | EPI_ISL_4252041 |
| COVM00653 | EPI_ISL_6094097 | COVC20860 | EPI_ISL_4252481 | KEM-21-03-95524 | EPI_ISL_4252045 |
| COVM00654 | EPI_ISL_6094100 | COVC23493 | EPI_ISL_4252482 | COVC15250 | EPI_ISL_4252046 |
| COVM00657 | EPI_ISL_6094115 | COVC11396 | EPI_ISL_4252483 | NHRL-S013 | EPI_ISL_4252050 |
| COVM00658 | EPI_ISL_6094120 | COVC21363 | EPI_ISL_4252484 | COVC20901 | EPI_ISL_4252053 |
| COVM00659 | EPI_ISL_6094121 | COVC14674 | EPI_ISL_4252485 | COVC14894 | EPI_ISL_4252055 |
| COVM00679 | EPI_ISL_6094147 | COVC15686 | EPI_ISL_4252492 | KEM-21-03-94090 | EPI_ISL_4252079 |
| COVM00681 | EPI_ISL_6094152 | COVC06252 | EPI_ISL_4252493 | COVC20819 | EPI_ISL_4252081 |
| COVM00691 | EPI_ISL_6094157 | COVC21005 | EPI_ISL_4252497 | COVC23508 | EPI_ISL_4252082 |
| COVM00692 | EPI_ISL_6094158 | COVC06031 | EPI_ISL_4252500 | COVC16131 | EPI_ISL_4252093 |
| COVM00693 | EPI_ISL_6094166 | COVC18558 | EPI_ISL_4252501 | COVC16213 | EPI_ISL_4252103 |
| COVM00702 | EPI_ISL_6094178 | COVC23894 | EPI_ISL_4252502 | KEM-21-03-95721 | EPI_ISL_4252114 |
| COVM00704 | EPI_ISL_6094179 | NHRL-S010 | EPI_ISL_4252504 | COVC08376 | EPI_ISL_4252110 |
| COVM00705 | EPI_ISL_6094185 | COVC23976 | EPI_ISL_4252505 | COVC16120 | EPI_ISL_4252113 |
| COVM00706 | EPI_ISL_6094192 | COVC15278 | EPI_ISL_4252509 | COVC04516 | EPI_ISL_4252115 |
| COVM00707 | EPI_ISL_6094199 | COVC21040 | EPI_ISL_4252513 | COVC24217 | EPI_ISL_4252125 |
| COVM00715 | EPI_ISL_6094215 | KEM-21-03-93916 | EPI_ISL_4252514 | COVC21008 | EPI_ISL_4252123 |
| COVM00721 | EPI_ISL_6094223 | COVC20900 | EPI_ISL_4252518 | NHRL-S024 | EPI_ISL_4252126 |
| COVM00723 | EPI_ISL_6094225 | COVC18572 | EPI_ISL_4252523 | NHRL-S032 | EPI_ISL_4252129 |
| COVM00724 | EPI_ISL_6094233 | COVC16085 | EPI_ISL_4252524 | COVC21037 | EPI_ISL_4252130 |

| | | | | | |
|---|---|---|---|---|---|
| COVM00727 | EPI_ISL_6094248 | COVC14699 | EPI_ISL_4252526 | COVC19917 | EPI_ISL_4252140 |
| COVM00728 | EPI_ISL_6094249 | COVC06454 | EPI_ISL_4252527 | COVC06226 | EPI_ISL_4252141 |
| COVM00736 | EPI_ISL_6094262 | COVC18712 | EPI_ISL_4252528 | KEM-21-03-95455 | EPI_ISL_4252143 |
| COVM00751 | EPI_ISL_6094299 | COVC21042 | EPI_ISL_4252530 | KEM-21-03-95565 | EPI_ISL_4252148 |
| COVM00753 | EPI_ISL_6094300 | COVC16216 | EPI_ISL_4252533 | COVC23715 | EPI_ISL_4252149 |
| COVM00754 | EPI_ISL_6094301 | COVC18658 | EPI_ISL_4252534 | COVC15008 | EPI_ISL_4252150 |
| COVM00755 | EPI_ISL_6094304 | KEM-21-03-94409 | EPI_ISL_4252538 | COVC14709 | EPI_ISL_4252151 |
| COVM00759 | EPI_ISL_6094306 | KEM-21-03-94654 | EPI_ISL_4252539 | COVC22775 | EPI_ISL_4252157 |
| COVM00770 | EPI_ISL_6094307 | KEM-21-03-93934 | EPI_ISL_4252540 | KEM-21-03-94683 | EPI_ISL_4252158 |
| COVM00773 | EPI_ISL_6094308 | KEM-21-03-95573 | EPI_ISL_4252543 | COVC16194 | EPI_ISL_4252170 |
| COVM00805 | EPI_ISL_6094333 | COVC23975 | EPI_ISL_4252548 | COVC16205 | EPI_ISL_4252171 |
| COVM00812 | EPI_ISL_6094344 | COVM00752 | EPI_ISL_8130691 | COVC16135 | EPI_ISL_4252177 |
| COVM00814 | EPI_ISL_6094353 | COVM00646 | EPI_ISL_8130671 | COVC21009 | EPI_ISL_4252178 |
| COVM00815 | EPI_ISL_6094355 | COVM01672 | EPI_ISL_8130710 | KEM-21-03-94676 | EPI_ISL_4252180 |
| COVM00826 | EPI_ISL_6094390 | COVM01684 | EPI_ISL_8130719 | COVC14947 | EPI_ISL_4252181 |
| COVM00841 | EPI_ISL_6094414 | COVM01675 | EPI_ISL_8130713 | NHRL-S026 | EPI_ISL_4252182 |
| COVM00843 | EPI_ISL_6094419 | COVM00689 | EPI_ISL_8130681 | COVC24032 | EPI_ISL_4252184 |
| COVM00844 | EPI_ISL_6094423 | COVM01678 | EPI_ISL_8130714 | COVC23670 | EPI_ISL_4252189 |
| COVM00845 | EPI_ISL_6094427 | COVM01679 | EPI_ISL_8130715 | COVC21420 | EPI_ISL_4252190 |
| COVM00847 | EPI_ISL_6094431 | COVM01680 | EPI_ISL_8130716 | NHRL-S035 | EPI_ISL_4252192 |
| COVM00848 | EPI_ISL_6094432 | COVM00764 | EPI_ISL_8130695 | COVC24215 | EPI_ISL_4252193 |
| COVM00850 | EPI_ISL_6094437 | COVM00680 | EPI_ISL_8130676 | COVC15114 | EPI_ISL_4252195 |
| COVM00851 | EPI_ISL_6094443 | COVM00648 | EPI_ISL_8130673 | COVC22829 | EPI_ISL_4252196 |
| COVM00852 | EPI_ISL_6094444 | COVM00686 | EPI_ISL_8130679 | KEM-21-03-95723 | EPI_ISL_4252204 |
| COVM00853 | EPI_ISL_6094448 | COVM00684 | EPI_ISL_8130678 | COVC23426 | EPI_ISL_4252315 |

| | | | | | |
|---|---|---|---|---|---|
| COVM00859 | EPI_ISL_6094456 | COVC26000 | EPI_ISL_8130670 | COVC23002 | EPI_ISL_4252206 |
| COVM00878 | EPI_ISL_6094493 | COVM00665 | EPI_ISL_8130675 | COVC23428 | EPI_ISL_4252209 |
| COVM00879 | EPI_ISL_6094497 | COVM01682 | EPI_ISL_8130718 | KEM-21-03-95585 | EPI_ISL_4252287 |
| COVM00884 | EPI_ISL_6094498 | COVM00698 | EPI_ISL_8130683 | COVC16210 | EPI_ISL_4252420 |
| COVM00916 | EPI_ISL_6094515 | COVM00763 | EPI_ISL_8130694 | COVC23438 | EPI_ISL_4252214 |
| COVM00944 | EPI_ISL_6094521 | COVM00830 | EPI_ISL_8130699 | COVC23534 | EPI_ISL_4252216 |
| COVM00945 | EPI_ISL_6094522 | COVM00683 | EPI_ISL_8130677 | COVC23550 | EPI_ISL_4252217 |
| COVM00975 | EPI_ISL_6094570 | COVM00696 | EPI_ISL_8130682 | COVC23705 | EPI_ISL_4252218 |
| COVM00976 | EPI_ISL_6094572 | COVM00711 | EPI_ISL_8130686 | KEM-21-03-93999 | EPI_ISL_4252221 |
| COVM00982 | EPI_ISL_6094579 | COVM00687 | EPI_ISL_8130680 | KEM-21-03-94005 | EPI_ISL_4252223 |
| COVM00993 | EPI_ISL_6094605 | COVM00701 | EPI_ISL_8130685 | KEM-21-03-94387 | EPI_ISL_4252224 |
| COVM00994 | EPI_ISL_6094606 | COVM00735 | EPI_ISL_8130689 | KEM-21-03-94640 | EPI_ISL_4252227 |
| COVM00996 | EPI_ISL_6094611 | COVM00730 | EPI_ISL_8130687 | KEM-21-03-95375 | EPI_ISL_4252233 |
| COVM00998 | EPI_ISL_6094613 | COVM00737 | EPI_ISL_8130690 | KEM-21-03-95522 | EPI_ISL_4252235 |
| COVM01004 | EPI_ISL_6094651 | COVM00734 | EPI_ISL_8130688 | COVC23635 | EPI_ISL_4252236 |
| COVM01005 | EPI_ISL_6094658 | COVM00757 | EPI_ISL_8130692 | KEM-21-03-93935 | EPI_ISL_4252239 |
| COVM01010 | EPI_ISL_6094659 | COVM00813 | EPI_ISL_8130698 | COVC23268 | EPI_ISL_4252240 |
| COVM01018 | EPI_ISL_6094688 | COVM00760 | EPI_ISL_8130693 | COVC23547 | EPI_ISL_4252241 |
| COVM01020 | EPI_ISL_6094693 | COVM00765 | EPI_ISL_8130696 | COVC23683 | EPI_ISL_4252242 |
| COVM01023 | EPI_ISL_6094697 | COVM00766 | EPI_ISL_8130697 | KEM-21-03-94389 | EPI_ISL_4252252 |
| COVM01024 | EPI_ISL_6094704 | COVM00660 | EPI_ISL_8130674 | KEM-21-03-94510 | EPI_ISL_4252249 |
| COVM01029 | EPI_ISL_6094719 | COVC25014 | EPI_ISL_8130636 | COVC18958 | EPI_ISL_4252253 |

| | | | | | |
|---|---|---|---|---|---|
| COVM01033 | EPI_ISL_6094727 | COVM00647 | EPI_ISL_8130672 | COVC21046 | EPI_ISL_4252261 |
| COVM01040 | EPI_ISL_6094732 | COVM01673 | EPI_ISL_8130711 | KEM-21-03-95517 | EPI_ISL_4252262 |
| COVM01044 | EPI_ISL_6094737 | COVM00700 | EPI_ISL_8130684 | KEM-21-03-95521 | EPI_ISL_4252264 |
| COVM01048 | EPI_ISL_6094743 | COVM01621 | EPI_ISL_9093416 | COVC18851 | EPI_ISL_4252265 |
| COVM01069 | EPI_ISL_6094754 | COVM01622 | EPI_ISL_9093417 | KEM-21-03-95688 | EPI_ISL_4252266 |
| COVC17732 | EPI_ISL_6095172 | COVM01619 | EPI_ISL_9093414 | COVC23911 | EPI_ISL_4252268 |
| COVC17742 | EPI_ISL_6095192 | COVM01620 | EPI_ISL_9093415 | KEM-21-03-94648 | EPI_ISL_4252269 |
| COVC17743 | EPI_ISL_6095200 | COVM01623 | EPI_ISL_9093418 | COVC03977 | EPI_ISL_4252271 |
| COVC17756 | EPI_ISL_6095217 | COVM01625 | EPI_ISL_9093419 | COVC19911 | EPI_ISL_4252272 |
| COVC17768 | EPI_ISL_6095222 | COVM01618 | EPI_ISL_9093413 | KEM-21-03-95703 | EPI_ISL_4252274 |
| COVC17774 | EPI_ISL_6095230 | COVM01636 | EPI_ISL_9093428 | COVC11236 | EPI_ISL_4252275 |
| COVC17776 | EPI_ISL_6095238 | COVM01628 | EPI_ISL_9093422 | COVC23593 | EPI_ISL_4252386 |
| COVC17792 | EPI_ISL_6095246 | COVM01635 | EPI_ISL_9093427 | KEM-21-03-95704 | EPI_ISL_4252281 |
| COVC17948 | EPI_ISL_6095328 | COVM01633 | EPI_ISL_9093425 | COVC19916 | EPI_ISL_4252283 |
| COVC17953 | EPI_ISL_6095332 | COVM01634 | EPI_ISL_9093426 | COVC21457 | EPI_ISL_4252284 |
| COVC17954 | EPI_ISL_6095333 | COVM01637 | EPI_ISL_9093429 | COVC16217 | EPI_ISL_4252506 |
| COVC17966 | EPI_ISL_6095338 | COVM01626 | EPI_ISL_9093420 | KEM-21-03-95553 | EPI_ISL_4252285 |
| COVC17979 | EPI_ISL_6095353 | COVM01642 | EPI_ISL_9093434 | KEM-21-03-95123 | EPI_ISL_4252288 |
| COVC17996 | EPI_ISL_6095356 | COVM01631 | EPI_ISL_9093423 | COVC20630 | EPI_ISL_4252295 |
| COVC17998 | EPI_ISL_6095363 | COVM01632 | EPI_ISL_9093424 | KEM-21-03-93949 | EPI_ISL_4252307 |
| COVC18025 | EPI_ISL_6095371 | COVM01627 | EPI_ISL_9093421 | COVC21036 | EPI_ISL_4252314 |
| COVC18051 | EPI_ISL_6095388 | COVM01646 | EPI_ISL_9093438 | COVC12936 | EPI_ISL_4252322 |

| | | | | | |
|---|---|---|---|---|---|
| COVC18082 | EPI_ISL_6095395 | COVM01644 | EPI_ISL_9093436 | COVC03813 | EPI_ISL_4252328 |
| COVC18104 | EPI_ISL_6095424 | COVM01645 | EPI_ISL_9093437 | COVC13749 | EPI_ISL_4252325 |
| COVC18161 | EPI_ISL_6095434 | COVM01638 | EPI_ISL_9093430 | COVC23957 | EPI_ISL_4252333 |
| COVC18190 | EPI_ISL_6095442 | COVM01639 | EPI_ISL_9093431 | COVC23886 | EPI_ISL_4252341 |
| COVC18192 | EPI_ISL_6095450 | COVM01643 | EPI_ISL_9093435 | COVC24019 | EPI_ISL_4252342 |
| COVC18287 | EPI_ISL_6095475 | COVM01640 | EPI_ISL_9093432 | KEM-21-03-95709 | EPI_ISL_4252344 |
| COVC18312 | EPI_ISL_6095490 | COVM01641 | EPI_ISL_9093433 | KEM-21-03-93925 | EPI_ISL_4252345 |
| COVC18325 | EPI_ISL_6095499 | COVC26044 | EPI_ISL_9093359 | COVC23267 | EPI_ISL_4252347 |
| COVC18350 | EPI_ISL_6095508 | COVC26041 | EPI_ISL_9093357 | KEM-21-03-95541 | EPI_ISL_4252348 |
| COVC18353 | EPI_ISL_6095519 | COVC26046 | EPI_ISL_9093360 | COVC23959 | EPI_ISL_4252349 |
| COVC18354 | EPI_ISL_6095525 | COVC26043 | EPI_ISL_9093358 | COVC03982 | EPI_ISL_4252352 |
| COVC18355 | EPI_ISL_6095535 | COVC26050 | EPI_ISL_9093363 | COVC23643 | EPI_ISL_4252357 |
| COVC18360 | EPI_ISL_6095546 | COVC24924 | EPI_ISL_9093356 | COVC03989 | EPI_ISL_4252358 |
| COVC18363 | EPI_ISL_6095554 | COVC26048 | EPI_ISL_9093361 | COVC23979 | EPI_ISL_4252363 |
| COVC18364 | EPI_ISL_6095555 | COVC26059 | EPI_ISL_9093364 | COVC14935 | EPI_ISL_4252364 |
| COVC18424 | EPI_ISL_6095556 | COVC26060 | EPI_ISL_9093365 | COVC16212 | EPI_ISL_4252365 |
| COVC18507 | EPI_ISL_6095567 | COVC26049 | EPI_ISL_9093362 | KEM-21-03-95237 | EPI_ISL_4252368 |
| COVC18512 | EPI_ISL_6095568 | COVC26068 | EPI_ISL_9093368 | COVC23928 | EPI_ISL_4252369 |
| COVC18524 | EPI_ISL_6095586 | COVC26069 | EPI_ISL_9093369 | COVC23951 | EPI_ISL_4252371 |
| COVC18526 | EPI_ISL_6095591 | COVC26074 | EPI_ISL_9093371 | COVC16150 | EPI_ISL_4252372 |
| COVC18527 | EPI_ISL_6095600 | COVC26061 | EPI_ISL_9093366 | COVC20839 | EPI_ISL_4252373 |
| COVC18570 | EPI_ISL_6095612 | COVC26072 | EPI_ISL_9093370 | COVC20689 | EPI_ISL_4252376 |
| COVC18587 | EPI_ISL_6095622 | COVC26065 | EPI_ISL_9093367 | COVC18989 | EPI_ISL_4252382 |
| COVC24352 | EPI_ISL_6095658 | COVM00459 | EPI_ISL_6093684 | COVC23568 | EPI_ISL_4252388 |
| COVC24355 | EPI_ISL_6095661 | COVM00461 | EPI_ISL_6093693 | COVC23575 | EPI_ISL_4252395 |
| COVC24471 | EPI_ISL_6095872 | COVM00464 | EPI_ISL_6093699 | COVC24003 | EPI_ISL_4252399 |

| | | | | | |
|---|---|---|---|---|---|
| COVM01179 | EPI_ISL_6095911 | COVM00467 | EPI_ISL_6093700 | COVC23453 | EPI_ISL_4252404 |
| COVM01184 | EPI_ISL_6095918 | COVM00479 | EPI_ISL_6093741 | COVC20862 | EPI_ISL_4252406 |
| COVM01186 | EPI_ISL_6095926 | COVM00483 | EPI_ISL_6093747 | COVC23744 | EPI_ISL_4252408 |
| COVM01199 | EPI_ISL_6095937 | COVM00484 | EPI_ISL_6093753 | COVC19804 | EPI_ISL_4252409 |
| COVM01207 | EPI_ISL_6095943 | COVM00490 | EPI_ISL_6093757 | COVC16174 | EPI_ISL_4252411 |
| COVM01211 | EPI_ISL_6095951 | COVM00528 | EPI_ISL_6093791 | COVC23206 | EPI_ISL_4252413 |
| COVM01212 | EPI_ISL_6095959 | COVM00503 | EPI_ISL_6093768 | COVC20865 | EPI_ISL_4252416 |
| COVM01219 | EPI_ISL_6095994 | COVM00551 | EPI_ISL_6093839 | COVC19908 | EPI_ISL_4252417 |
| COVM01225 | EPI_ISL_6096009 | COVM00555 | EPI_ISL_6093847 | KEM-21-03-95591 | EPI_ISL_4252418 |
| COVM01230 | EPI_ISL_6096016 | COVM00562 | EPI_ISL_6093879 | COVC13800 | EPI_ISL_4252423 |
| COVM01231 | EPI_ISL_6096022 | COVM00572 | EPI_ISL_6093914 | KEM-21-03-95540 | EPI_ISL_4252425 |
| COVM01238 | EPI_ISL_6096034 | COVM00580 | EPI_ISL_6093944 | COVC21391 | EPI_ISL_4252430 |
| COVM01239 | EPI_ISL_6096041 | COVM00585 | EPI_ISL_6093965 | COVC23637 | EPI_ISL_4252431 |
| COVM01240 | EPI_ISL_6096049 | COVM00586 | EPI_ISL_6093970 | COVC19790 | EPI_ISL_4252434 |
| COVM01241 | EPI_ISL_6096052 | COVM00594 | EPI_ISL_6093984 | COVC15674 | EPI_ISL_4252435 |
| COVM01257 | EPI_ISL_6096083 | COVM00595 | EPI_ISL_6093987 | COVC23974 | EPI_ISL_4252439 |
| COVM01259 | EPI_ISL_6096084 | COVM00599 | EPI_ISL_6093996 | COVC06232 | EPI_ISL_4252441 |
| COVM01264 | EPI_ISL_6096085 | COVM00607 | EPI_ISL_6094006 | COVC24169 | EPI_ISL_4252442 |
| COVM01272 | EPI_ISL_6096099 | COVM00608 | EPI_ISL_6094012 | COVC12481 | EPI_ISL_4252443 |
| COVM01275 | EPI_ISL_6096102 | COVM00619 | EPI_ISL_6094044 | KEM-21-03-94684 | EPI_ISL_4252445 |
| COVM01282 | EPI_ISL_6096134 | COVM00621 | EPI_ISL_6094048 | COVC19011 | EPI_ISL_4252449 |
| COVM01283 | EPI_ISL_6096143 | COVM00622 | EPI_ISL_6094049 | COVC24072 | EPI_ISL_4252450 |
| COVM01284 | EPI_ISL_6096152 | COVM00624 | EPI_ISL_6094055 | COVC22965 | EPI_ISL_4252454 |
| COVM01290 | EPI_ISL_6096181 | COVM00630 | EPI_ISL_6094063 | KEM-21-03-95587 | EPI_ISL_4252455 |
| COVM01292 | EPI_ISL_6096184 | COVM00633 | EPI_ISL_6094069 | COVC21447 | EPI_ISL_4252462 |
| COVM01293 | EPI_ISL_6096189 | COVM00644 | EPI_ISL_6094075 | COVC23440 | EPI_ISL_4252463 |

| | | | | | |
|---|---|---|---|---|---|
| COVM01300 | EPI_ISL_6096224 | COVM00655 | EPI_ISL_6094108 | COVC23506 | EPI_ISL_4252467 |
| COVM01301 | EPI_ISL_6096225 | COVM00662 | EPI_ISL_6094127 | COVC20856 | EPI_ISL_4252469 |
| COVM01303 | EPI_ISL_6096226 | COVM00663 | EPI_ISL_6094133 | COVC23977 | EPI_ISL_4252544 |
| COVM01310 | EPI_ISL_6096335 | COVM00673 | EPI_ISL_6094139 | COVC20770 | EPI_ISL_4252472 |
| COVM01314 | EPI_ISL_6096344 | COVM00675 | EPI_ISL_6094140 | COVC15026 | EPI_ISL_4252476 |
| COVM01317 | EPI_ISL_6096347 | COVM00697 | EPI_ISL_6094169 | COVC21464 | EPI_ISL_4252479 |
| COVM01320 | EPI_ISL_6096351 | COVM00699 | EPI_ISL_6094173 | COVC14794 | EPI_ISL_4252486 |
| COVM01326 | EPI_ISL_6096403 | COVM00710 | EPI_ISL_6094204 | KEM-21-03-94667 | EPI_ISL_4252488 |
| COVM01328 | EPI_ISL_6096408 | COVM00712 | EPI_ISL_6094205 | COVC16128 | EPI_ISL_4252489 |
| COVM01329 | EPI_ISL_6096412 | COVM00713 | EPI_ISL_6094209 | COVC16186 | EPI_ISL_4252490 |
| COVM01337 | EPI_ISL_6096428 | COVM00716 | EPI_ISL_6094221 | COVC16091 | EPI_ISL_4252491 |
| COVM01347 | EPI_ISL_6096435 | COVM00725 | EPI_ISL_6094243 | COVC14676 | EPI_ISL_4252496 |
| COVM01351 | EPI_ISL_6096436 | COVM00792 | EPI_ISL_6094319 | COVC14793 | EPI_ISL_4252498 |
| COVM01352 | EPI_ISL_6096444 | COVM00733 | EPI_ISL_6094258 | NHRL-S031 | EPI_ISL_4252511 |
| COVM01358 | EPI_ISL_6096467 | COVM00746 | EPI_ISL_6094288 | COVC23498 | EPI_ISL_4252515 |
| COVM01362 | EPI_ISL_6096482 | COVM00738 | EPI_ISL_6094271 | COVC06233 | EPI_ISL_4252516 |
| COVM01364 | EPI_ISL_6096489 | COVM00739 | EPI_ISL_6094278 | COVC19789 | EPI_ISL_4252517 |
| COVM01365 | EPI_ISL_6096490 | COVM00748 | EPI_ISL_6094289 | COVC08344 | EPI_ISL_4252522 |
| COVM01374 | EPI_ISL_6096504 | COVM00758 | EPI_ISL_6094305 | COVC20459 | EPI_ISL_4252529 |
| COVM01375 | EPI_ISL_6096506 | COVM00791 | EPI_ISL_6094314 | COVC16155 | EPI_ISL_4252532 |
| COVM01377 | EPI_ISL_6096511 | COVM00803 | EPI_ISL_6094325 | COVC20629 | EPI_ISL_4252537 |
| COVM01380 | EPI_ISL_6096516 | COVM00804 | EPI_ISL_6094326 | COVC24146 | EPI_ISL_4252541 |
| COVM01388 | EPI_ISL_6096560 | COVM00811 | EPI_ISL_6094340 | KEM-21-03-94499 | EPI_ISL_4252545 |
| COVM01398 | EPI_ISL_6096587 | COVM00816 | EPI_ISL_6094362 | COVC06059 | EPI_ISL_4252546 |
| COVM01400 | EPI_ISL_6096589 | COVM00820 | EPI_ISL_6094366 | COVC21077 | EPI_ISL_4252547 |
| COVM01402 | EPI_ISL_6096592 | COVM00821 | EPI_ISL_6094367 | COVC23362 | EPI_ISL_4252550 |
| COVM01406 | EPI_ISL_6096602 | COVM00822 | EPI_ISL_6094377 | COVC24026 | EPI_ISL_4252551 |

| | | | | | |
|---|---|---|---|---|---|
| COVM01408 | EPI_ISL_6096609 | COVM00823 | EPI_ISL_6094383 | COVC20767 | EPI_ISL_4252553 |
| COVM01409 | EPI_ISL_6096610 | COVM00825 | EPI_ISL_6094384 | COVM01647 | EPI_ISL_9093439 |
| COVM01413 | EPI_ISL_6096622 | COVM00842 | EPI_ISL_6094418 | COVM01648 | EPI_ISL_9093440 |
| COVM01417 | EPI_ISL_6096628 | COVM00827 | EPI_ISL_6094393 | COVM01649 | EPI_ISL_9093441 |
| COVM00992 | EPI_ISL_6096663 | COVM00828 | EPI_ISL_6094399 | COVM01671 | EPI_ISL_9093442 |
| COVM01052 | EPI_ISL_6096670 | COVM00832 | EPI_ISL_6094400 | COVM01676 | EPI_ISL_9093443 |
| COVC17912 | EPI_ISL_6096767 | COVM00835 | EPI_ISL_6094408 | COVM01677 | EPI_ISL_9093444 |
| COVC17935 | EPI_ISL_6096777 | COVM01002 | EPI_ISL_6094639 | COVM01683 | EPI_ISL_9093445 |
| COVC18017 | EPI_ISL_6096786 | COVM00860 | EPI_ISL_6094462 | COVM01685 | EPI_ISL_9093446 |
| COVM01188 | EPI_ISL_6096823 | COVM00865 | EPI_ISL_6094463 | COVM01696 | EPI_ISL_9093456 |
| COVM01205 | EPI_ISL_6096826 | COVM00873 | EPI_ISL_6094477 | COVM01698 | EPI_ISL_9093457 |
| COVM01228 | EPI_ISL_6096830 | COVM00867 | EPI_ISL_6094469 | COVM01699 | EPI_ISL_9093458 |
| COVM01390 | EPI_ISL_6096831 | COVM00874 | EPI_ISL_6094485 | COVM01700 | EPI_ISL_9093459 |
| COVC03617 | EPI_ISL_4251927 | COVM00885 | EPI_ISL_6094503 | COVM01701 | EPI_ISL_9093460 |
| COVC03696 | EPI_ISL_4251928 | COVM00887 | EPI_ISL_6094508 | COVM01702 | EPI_ISL_9093461 |
| COVC04189 | EPI_ISL_4251933 | COVM00951 | EPI_ISL_6094530 | COVM01703 | EPI_ISL_9093462 |
| COVC04192 | EPI_ISL_4251934 | COVM00952 | EPI_ISL_6094536 | COVM01704 | EPI_ISL_9093463 |
| COVC07861 | EPI_ISL_4251936 | COVM00953 | EPI_ISL_6094540 | COVM01705 | EPI_ISL_9093464 |
| COVC08637 | EPI_ISL_4251937 | COVM00956 | EPI_ISL_6094550 | COVM01706 | EPI_ISL_9093465 |
| COVC11223 | EPI_ISL_4251938 | COVM00960 | EPI_ISL_6094555 | COVM01707 | EPI_ISL_9093466 |
| COVC11243 | EPI_ISL_4251939 | COVM00961 | EPI_ISL_6094556 | COVM01708 | EPI_ISL_9093467 |
| COVC12478 | EPI_ISL_4251941 | COVM00963 | EPI_ISL_6094561 | COVM01709 | EPI_ISL_9093468 |
| COVC18970 | EPI_ISL_4251948 | COVM00983 | EPI_ISL_6094580 | COVM01710 | EPI_ISL_9093469 |
| COVC18990 | EPI_ISL_4251949 | COVM00985 | EPI_ISL_6094589 | COVM01744 | EPI_ISL_9093495 |
| COVC20543 | EPI_ISL_4251950 | COVM00990 | EPI_ISL_6094598 | COVM01746 | EPI_ISL_9093496 |
| COVC21060 | EPI_ISL_4251952 | COVM01000 | EPI_ISL_6094620 | COVM01748 | EPI_ISL_9093497 |
| COVC21062 | EPI_ISL_4251953 | COVM01001 | EPI_ISL_6094630 | COVM01749 | EPI_ISL_9093498 |
| COVC21166 | EPI_ISL_4251954 | COVM01003 | EPI_ISL_6094641 | COVM01750 | EPI_ISL_9093499 |
| COVC21327 | EPI_ISL_4251956 | COVM01015 | EPI_ISL_6094669 | COVM01753 | EPI_ISL_9093500 |

| | | | | | |
|---|---|---|---|---|---|
| COVC23639 | EPI_ISL_4251957 | COVM01016 | EPI_ISL_6094670 | COVM01754 | EPI_ISL_9093501 |
| COVC20807 | EPI_ISL_4251962 | COVM01017 | EPI_ISL_6094676 | COVM01755 | EPI_ISL_9093502 |
| COVC20844 | EPI_ISL_4251963 | COVM01026 | EPI_ISL_6094712 | COVM01756 | EPI_ISL_9093503 |
| COVC20854 | EPI_ISL_4251964 | COVC17735 | EPI_ISL_6095184 | COVM01757 | EPI_ISL_9093504 |
| COVC20864 | EPI_ISL_4251965 | COVM01050 | EPI_ISL_6094748 | COVM01758 | EPI_ISL_9093505 |
| COVC21470 | EPI_ISL_4251966 | COVC17734 | EPI_ISL_6095178 | COVM01759 | EPI_ISL_9093506 |
| COVC20855 | EPI_ISL_4251971 | COVC17739 | EPI_ISL_6095185 | COVM01760 | EPI_ISL_9093507 |
| COVC23988 | EPI_ISL_4251973 | COVC17740 | EPI_ISL_6095186 | COVM01761 | EPI_ISL_9093508 |
| COVC24148 | EPI_ISL_4251974 | COVC17750 | EPI_ISL_6095206 | COVM01762 | EPI_ISL_9093509 |
| COVC23887 | EPI_ISL_4251988 | COVC17810 | EPI_ISL_6095250 | COVM01763 | EPI_ISL_9093510 |
| NHRL-S028 | EPI_ISL_4251990 | COVC17854 | EPI_ISL_6095260 | COVM01764 | EPI_ISL_9093511 |
| COVC21003 | EPI_ISL_4251991 | COVC17858 | EPI_ISL_6095269 | COVM01766 | EPI_ISL_9093513 |
| COVC23589 | EPI_ISL_4251994 | COVC17916 | EPI_ISL_6095279 | COVM01767 | EPI_ISL_9093514 |
| KEM-21-03-95590 | EPI_ISL_4251995 | COVC17919 | EPI_ISL_6095286 | COVM01768 | EPI_ISL_9093515 |
| COVC23497 | EPI_ISL_4251999 | COVC17920 | EPI_ISL_6095292 | COVM01769 | EPI_ISL_9093516 |
| COVC23006 | EPI_ISL_4252001 | COVC17922 | EPI_ISL_6095301 | COVM01771 | EPI_ISL_9093518 |
| COVC22920 | EPI_ISL_4252005 | COVC17923 | EPI_ISL_6095308 | COVM01799 | EPI_ISL_9093539 |
| COVC19990 | EPI_ISL_4252006 | COVC17924 | EPI_ISL_6095317 | COVM01800 | EPI_ISL_9093540 |
| COVC06225 | EPI_ISL_4252008 | COVC17946 | EPI_ISL_6095324 | COVM01811 | EPI_ISL_9093551 |
| NHRL-S012 | EPI_ISL_4252009 | COVC17978 | EPI_ISL_6095345 | COVM00469 | EPI_ISL_9746936 |
| COVC22975 | EPI_ISL_4252011 | COVC18032 | EPI_ISL_6095380 | COVC17771 | EPI_ISL_9746937 |
| COVC23641 | EPI_ISL_4252012 | COVC18093 | EPI_ISL_6095401 | COVC17747 | EPI_ISL_9746938 |
| COVC20861 | EPI_ISL_4252017 | COVC18097 | EPI_ISL_6095409 | COVM00587 | EPI_ISL_9746939 |
| COVC22972 | EPI_ISL_4252018 | COVC18101 | EPI_ISL_6095417 | COVM01009 | EPI_ISL_9746940 |
| COVC23354 | EPI_ISL_4252019 | COVC18210 | EPI_ISL_6095459 | COVM01175 | EPI_ISL_9746941 |
| COVC20640 | EPI_ISL_4252020 | COVC18286 | EPI_ISL_6095469 | COVM00641 | EPI_ISL_9746943 |
| COVC21483 | EPI_ISL_4252022 | COVC18288 | EPI_ISL_6095476 | COVC18605 | EPI_ISL_9746944 |
| COVC16029 | EPI_ISL_4252028 | COVC18295 | EPI_ISL_6095483 | COVM00866 | EPI_ISL_9746945 |
| COVC20622 | EPI_ISL_4252030 | COVC18497 | EPI_ISL_6095563 | COVM00980 | EPI_ISL_9746946 |

| | | | | | |
|---|---|---|---|---|---|
| COVC20619 | EPI_ISL_4252033 | COVC18516 | EPI_ISL_6095569 | COVM00868 | EPI_ISL_9746947 |
| COVC23968 | EPI_ISL_4252034 | COVC18518 | EPI_ISL_6095577 | COVM00987 | EPI_ISL_9746948 |
| KEM-21-03-94007 | EPI_ISL_4252035 | COVM01194 | EPI_ISL_6095929 | COVM00573 | EPI_ISL_9746950 |
| COVC23689 | EPI_ISL_4252038 | COVC18520 | EPI_ISL_6095582 | COVC18015 | EPI_ISL_9746951 |
| KEM-21-03-94736 | EPI_ISL_4252039 | COVM01215 | EPI_ISL_6095976 | COVM01360 | EPI_ISL_9746952 |
| KEM-21-03-93919 | EPI_ISL_4252040 | COVC18559 | EPI_ISL_6095607 | COVM01336 | EPI_ISL_9746953 |
| COVC16214 | EPI_ISL_4252047 | COVC18592 | EPI_ISL_6095630 | COVM01189 | EPI_ISL_9746954 |
| COVC23190 | EPI_ISL_4252049 | COVC18597 | EPI_ISL_6095636 | COVM00719 | EPI_ISL_9746955 |
| COVC21315 | EPI_ISL_4252051 | COVM01245 | EPI_ISL_6096061 | COVM01311 | EPI_ISL_9746958 |
| COVC14948 | EPI_ISL_4252052 | COVC24472 | EPI_ISL_6095882 | COVM01363 | EPI_ISL_9746959 |
| COVC23579 | EPI_ISL_4252060 | COVC24474 | EPI_ISL_6095886 | COVM01182 | EPI_ISL_9746960 |
| COVC23592 | EPI_ISL_4252063 | COVM01176 | EPI_ISL_6095893 | COVM00629 | EPI_ISL_9746961 |
| NHRL-S001 | EPI_ISL_4252064 | COVM01177 | EPI_ISL_6095902 | COVM00995 | EPI_ISL_9746962 |
| COVC23189 | EPI_ISL_4252065 | COVM01191 | EPI_ISL_6095927 | COVM00537 | EPI_ISL_9746963 |
| COVC21432 | EPI_ISL_4252066 | COVM01193 | EPI_ISL_6095928 | COVM00779 | EPI_ISL_9746964 |
| COVC21032 | EPI_ISL_4252068 | COVM01214 | EPI_ISL_6095969 | COVM00536 | EPI_ISL_9746965 |
| KEM-21-03-95564 | EPI_ISL_4252069 | COVM01217 | EPI_ISL_6095984 | COVM00717 | EPI_ISL_9746966 |
| COVC11266 | EPI_ISL_4252070 | COVM01223 | EPI_ISL_6096001 | COVM00578 | EPI_ISL_9746967 |
| COVC24132 | EPI_ISL_4252072 | COVM01233 | EPI_ISL_6096028 | COVM00605 | EPI_ISL_9746969 |
| NHRL-S011 | EPI_ISL_4252073 | COVM01304 | EPI_ISL_6096231 | COVM00569 | EPI_ISL_9746970 |
| NHRL-S027 | EPI_ISL_4252074 | COVM01249 | EPI_ISL_6096066 | COVC17737 | EPI_ISL_9746971 |
| COVC22969 | EPI_ISL_4252075 | COVM01252 | EPI_ISL_6096073 | COVM00477 | EPI_ISL_9746972 |
| COVC14753 | EPI_ISL_4252077 | COVM01265 | EPI_ISL_6096086 | COVC17921 | EPI_ISL_9746973 |
| KEM-21-03-94519 | EPI_ISL_4252078 | COVM01270 | EPI_ISL_6096088 | COVM01187 | EPI_ISL_9746974 |
| COVC22934 | EPI_ISL_4252080 | COVM01278 | EPI_ISL_6096112 | COVM00511 | EPI_ISL_9746975 |
| COVC20831 | EPI_ISL_4252084 | COVM01280 | EPI_ISL_6096121 | COVM01210 | EPI_ISL_9746976 |
| COVC14784 | EPI_ISL_4252085 | COVM01281 | EPI_ISL_6096129 | COVM00478 | EPI_ISL_9746979 |
| COVC24216 | EPI_ISL_4252086 | COVM01286 | EPI_ISL_6096156 | COVC17784 | EPI_ISL_9746980 |
| COVC23925 | EPI_ISL_4252087 | COVM01287 | EPI_ISL_6096166 | COVM00557 | EPI_ISL_9746982 |

| | | | | | |
|---|---|---|---|---|---|
| COVC21456 | EPI_ISL_4252089 | COVM01288 | EPI_ISL_6096171 | COVM01253 | EPI_ISL_9746983 |
| COVC19955 | EPI_ISL_4252090 | COVM01294 | EPI_ISL_6096195 | COVM00488 | EPI_ISL_9746985 |
| COVC21328 | EPI_ISL_4252091 | COVM01296 | EPI_ISL_6096200 | COVM01051 | EPI_ISL_9746987 |
| KEM-21-03-93941 | EPI_ISL_4252092 | COVM01297 | EPI_ISL_6096207 | COVM00489 | EPI_ISL_9746988 |
| COVC06654 | EPI_ISL_4252094 | COVM01298 | EPI_ISL_6096212 | COVM00954 | EPI_ISL_9746990 |
| COVC18941 | EPI_ISL_4252095 | COVM01299 | EPI_ISL_6096222 | COVM01313 | EPI_ISL_9746991 |
| COVC18673 | EPI_ISL_4252096 | COVM01305 | EPI_ISL_6096240 | COVM00559 | EPI_ISL_9746992 |
| COVC22801 | EPI_ISL_4252097 | COVM01307 | EPI_ISL_6096247 | COVM00973 | EPI_ISL_9746993 |
| COVC06246 | EPI_ISL_4252099 | COVM01308 | EPI_ISL_6096256 | COVM01277 | EPI_ISL_9746994 |
| COVC11235 | EPI_ISL_4252100 | COVM01321 | EPI_ISL_6096360 | COVM01046 | EPI_ISL_9746995 |
| COVC18975 | EPI_ISL_4252102 | COVM01322 | EPI_ISL_6096367 | COVC17757 | EPI_ISL_9746996 |
| COVC24014 | EPI_ISL_4252104 | COVM01323 | EPI_ISL_6096375 | COVM01372 | EPI_ISL_9746998 |
| COVC16116 | EPI_ISL_4252105 | COVM01324 | EPI_ISL_6096384 | COVM00840 | EPI_ISL_9747002 |
| COVC06200 | EPI_ISL_4252109 | COVM01325 | EPI_ISL_6096393 | COVM00678 | EPI_ISL_9747003 |
| COVC20899 | EPI_ISL_4252111 | COVM01333 | EPI_ISL_6096415 | COVM00802 | EPI_ISL_9747005 |
| COVC15059 | EPI_ISL_4252112 | COVM01335 | EPI_ISL_6096424 | COVM00749 | EPI_ISL_9747006 |
| COVC04519 | EPI_ISL_4252116 | COVM01339 | EPI_ISL_6096429 | COVM00750 | EPI_ISL_9747007 |
| NHRL-S036 | EPI_ISL_4252117 | COVM01340 | EPI_ISL_6096430 | COVM00685 | EPI_ISL_9749493 |
| COVC16188 | EPI_ISL_4252120 | COVM01341 | EPI_ISL_6096431 | COVM00768 | EPI_ISL_9749494 |
| NHRL-S029 | EPI_ISL_4252122 | COVM01342 | EPI_ISL_6096432 | COVM01745 | EPI_ISL_9749498 |
| KEM-21-03-94421 | EPI_ISL_4252131 | COVM01345 | EPI_ISL_6096434 | COVM01752 | EPI_ISL_9749499 |
| KEM-21-03-95536 | EPI_ISL_4252132 | COVM01353 | EPI_ISL_6096450 | KEMRI_01_89637 | EPI_ISL_4251960 |
| COVC20893 | EPI_ISL_4252133 | COVM01354 | EPI_ISL_6096452 | KEMRI_02_92670 | EPI_ISL_4252029 |
| KEM-21-03-93899 | EPI_ISL_4252134 | COVM01355 | EPI_ISL_6096456 | KEMRI_02_93645 | EPI_ISL_4252056 |
| COVC19995 | EPI_ISL_4252135 | COVM01356 | EPI_ISL_6096463 | KEMRI_02_91769 | EPI_ISL_4252057 |

| | | | | | |
|---|---|---|---|---|---|
| COVC13805 | EPI_ISL_4252136 | COVM01357 | EPI_ISL_6096466 | KEMRI_01_89447 | EPI_ISL_4252067 |
| KEM-21-03-94701 | EPI_ISL_4252137 | COVM01359 | EPI_ISL_6096473 | KEMRI_01_89476 | EPI_ISL_4252071 |
| COVC21011 | EPI_ISL_4252138 | COVM01361 | EPI_ISL_6096476 | KEMRI_02_93806 | EPI_ISL_4252098 |
| KEM-21-03-93945 | EPI_ISL_4252142 | COVM01366 | EPI_ISL_6096495 | KEMRI_02_91762 | EPI_ISL_4252106 |
| COVC04527 | EPI_ISL_4252144 | COVM01368 | EPI_ISL_6096498 | KEMRI_01_91179 | EPI_ISL_4252119 |
| COVC20649 | EPI_ISL_4252145 | COVM01373 | EPI_ISL_6096503 | KEMRI_01_91178 | EPI_ISL_4252128 |
| COVC23352 | EPI_ISL_4252152 | COVM01379 | EPI_ISL_6096515 | KEMRI_02_91743 | EPI_ISL_4252200 |
| COVC24068 | EPI_ISL_4252153 | COVM01381 | EPI_ISL_6096523 | KEMRI_02_92896 | EPI_ISL_4252254 |
| COVC23712 | EPI_ISL_4252154 | COVM01382 | EPI_ISL_6096526 | KEMRI_02_93544 | EPI_ISL_4252267 |
| KEM-21-03-94425 | EPI_ISL_4252155 | COVM01383 | EPI_ISL_6096534 | KEMRI_02_93550 | EPI_ISL_4252379 |
| COVC08336 | EPI_ISL_4252156 | COVM01385 | EPI_ISL_6096542 | KEMRI_02_93270 | EPI_ISL_4252385 |
| COVC15422 | EPI_ISL_4252159 | COVM01387 | EPI_ISL_6096551 | KEMRI_01_89767 | EPI_ISL_4252387 |
| COVC21024 | EPI_ISL_4252160 | COVM01403 | EPI_ISL_6096594 | KEMRI_02_92305 | EPI_ISL_4252392 |
| COVC15036 | EPI_ISL_4252161 | COVM01411 | EPI_ISL_6096620 | KEMRI_01_90285 | EPI_ISL_4252394 |
| KEM-21-03-95594 | EPI_ISL_4252162 | COVM01412 | EPI_ISL_6096621 | KEMRI_01_89768 | EPI_ISL_4252448 |
| KEM-21-03-95580 | EPI_ISL_4252163 | COVM01416 | EPI_ISL_6096623 | KEMRI_01_89581 | EPI_ISL_4252466 |
| COVC20840 | EPI_ISL_4252164 | COVM01065 | EPI_ISL_6096671 | KEMRI_02_91761 | EPI_ISL_4252468 |

| | | | | | |
|---|---|---|---|---|---|
| COVC21405 | EPI_ISL_4252167 | COVC17910 | EPI_ISL_6096766 | KEMRI_02_93273 | EPI_ISL_4252494 |
| COVC23472 | EPI_ISL_4252168 | KEM-21-03-94511 | EPI_ISL_8318522 | KEMRI_02_91737 | EPI_ISL_4252520 |
| COVC22971 | EPI_ISL_4252169 | KEM-21-03-95515 | EPI_ISL_8318521 | KEMRI_01_89487 | EPI_ISL_4252531 |
| COVC15001 | EPI_ISL_4252172 | COVC21398 | EPI_ISL_8318532 | KEMRI_02_92563 | EPI_ISL_8318515 |
| COVC20768 | EPI_ISL_4252176 | COVC24007 | EPI_ISL_8318533 | KEMRI_02_92681 | EPI_ISL_8318517 |
| COVC22973 | EPI_ISL_4252179 | COVC23198 | EPI_ISL_8318536 | KEMRI_02_91770 | EPI_ISL_8318518 |
| COVC23685 | EPI_ISL_4252183 | COVC14801 | EPI_ISL_8318547 | KEMRI_02_92570 | EPI_ISL_8318526 |
| COVC18853 | EPI_ISL_4252185 | COVC24058 | EPI_ISL_8318546 | KEMRI_02_91764 | EPI_ISL_8318531 |
| NHRL-S037 | EPI_ISL_4252186 | COVC23477 | EPI_ISL_8318575 | KEMRI_02_92441 | EPI_ISL_8318538 |
| COVC20826 | EPI_ISL_4252187 | COVC19336 | EPI_ISL_8318604 | KEMRI_01_91165 | EPI_ISL_8318561 |
| COVC18546 | EPI_ISL_4252188 | COVC23999 | EPI_ISL_8318613 | KEMRI_01_89716 | EPI_ISL_8318563 |
| COVC24145 | EPI_ISL_4252191 | KEM-21-03-94502 | EPI_ISL_8318594 | KEMRI_02_93127 | EPI_ISL_8318564 |
| COVC23197 | EPI_ISL_4252198 | COVC16191 | EPI_ISL_8318596 | KEMRI_01_90728 | EPI_ISL_8318570 |
| COVC04209 | EPI_ISL_4252201 | COVC23552 | EPI_ISL_8318592 | KEMRI_01_91176 | EPI_ISL_8318577 |
| COVC15255 | EPI_ISL_4252202 | COVC23353 | EPI_ISL_8318590 | KEMRI_02_92704 | EPI_ISL_8318582 |
| COVC21475 | EPI_ISL_4252203 | COVC23572 | EPI_ISL_8318574 | KEMRI_01_91183 | EPI_ISL_8318588 |
| COVC21058 | EPI_ISL_4252205 | COVC23473 | EPI_ISL_8318576 | KEMRI_02_91731 | EPI_ISL_8318589 |

| | | | | | |
|---|---|---|---|---|---|
| COVC23313 | EPI_ISL_4252207 | COVC23611 | EPI_ISL_8318578 | KEMRI_01_91173 | EPI_ISL_8318591 |
| COVC23355 | EPI_ISL_4252208 | COVC23634 | EPI_ISL_8318572 | KEMRI_02_93271 | EPI_ISL_8318593 |
| COVC23429 | EPI_ISL_4252210 | COVC23917 | EPI_ISL_8318579 | KEMRI_02_91757 | EPI_ISL_8318595 |
| COVC23430 | EPI_ISL_4252211 | KEM-21-03-93946 | EPI_ISL_8318584 | KEMRI_02_93781 | EPI_ISL_8318598 |
| COVC23431 | EPI_ISL_4252212 | COVC23743 | EPI_ISL_8318586 | KEMRI_02_91728 | EPI_ISL_8318599 |
| COVC23433 | EPI_ISL_4252213 | COVC23989 | EPI_ISL_8318585 | KEMRI_02_92919 | EPI_ISL_8318600 |
| COVC23450 | EPI_ISL_4252215 | COVC23669 | EPI_ISL_8318583 | KEMRI_02_91742 | EPI_ISL_8318602 |
| KEM-21-03-93738 | EPI_ISL_4252220 | KEM-21-03-94650 | EPI_ISL_8318581 | KEMRI_02_91739 | EPI_ISL_8318603 |
| KEM-21-03-94000 | EPI_ISL_4252222 | COVC23490 | EPI_ISL_8318580 | KEMRI_02_91745 | EPI_ISL_8318606 |
| KEM-21-03-94388 | EPI_ISL_4252225 | COVC23437 | EPI_ISL_8318566 | KEMRI_01_90725 | EPI_ISL_8318615 |
| KEM-21-03-94632 | EPI_ISL_4252226 | COVC14892 | EPI_ISL_8318553 | COVC20830 | EPI_ISL_4252305 |
| KEM-21-03-94644 | EPI_ISL_4252228 | COVC23193 | EPI_ISL_8318558 | KEM-21-03-94675 | EPI_ISL_4252306 |
| KEM-21-03-94710 | EPI_ISL_4252229 | COVC19668 | EPI_ISL_4252303 | COVC23932 | EPI_ISL_4252308 |
| KEM-21-03-95121 | EPI_ISL_4252230 | KEM-21-03-94709 | EPI_ISL_4252256 | COVC20842 | EPI_ISL_4252286 |
| KEM-21-03-95255 | EPI_ISL_4252231 | COVC04150 | EPI_ISL_4252257 | COVC19824 | EPI_ISL_4252290 |
| KEM-21-03-95366 | EPI_ISL_4252232 | KEM-21-03-95520 | EPI_ISL_4252263 | COVC19030 | EPI_ISL_4252292 |
| KEM-21-03-95519 | EPI_ISL_4252234 | KEM-21-03-95534 | EPI_ISL_4252270 | COVC23441 | EPI_ISL_4252293 |
| COVC23754 | EPI_ISL_4252243 | COVC23347 | EPI_ISL_4252276 | COVC20845 | EPI_ISL_4252297 |
| KEM-21-03-94498 | EPI_ISL_4252248 | COVC20835 | EPI_ISL_4252277 | COVC23706 | EPI_ISL_4252298 |
| NHRL-S003 | EPI_ISL_4252250 | KEM-21-03-95708 | EPI_ISL_4252279 | COVC23548 | EPI_ISL_4252300 |
| COVC23686 | EPI_ISL_4252251 | COVC20827 | EPI_ISL_4252282 | COVC23915 | EPI_ISL_4252302 |

| **Supplement Table 2.** Accession Numbers for SARS-CoV genomes submitted to NCBI | | | |
|---|---|---|---|
| **Local Name** | **Accession Number** | **Local Name** | **Accession Number** |
| ILRI_COVC14964 | OM738210 | ILRI_COVC24147 | OM738240 |
| ILRI_COVC15112 | OM738211 | ILRI_COVC24473 | OM738241 |
| ILRI_COVC15443 | OM738212 | ILRI_COVM00545 | OM738242 |
| ILRI_COVC15641 | OM738213 | ILRI_COVM00550 | OM738243 |
| ILRI_COVC15679 | OM738214 | ILRI_COVM00557 | OM738244 |
| ILRI_COVC16034 | OM738215 | ILRI_COVM00694 | OM738245 |
| ILRI_COVC16148 | OM738216 | ILRI_COVM01022 | OM738246 |
| ILRI_COVC16151 | OM738217 | ILRI_COVM01049 | OM738247 |
| ILRI_COVC16203 | OM738218 | ILRI_COVM01180 | OM738248 |
| ILRI_COVC17903 | OM738219 | ILRI_COVM01190 | OM738249 |
| ILRI_COVC17983 | OM738220 | ILRI_COVM01192 | OM738250 |
| ILRI_COVC18434 | OM738221 | ILRI_COVM01195 | OM738251 |
| ILRI_COVC18439 | OM738222 | ILRI_COVM01196 | OM738252 |
| ILRI_COVC18443 | OM738223 | ILRI_COVM01197 | OM738253 |
| ILRI_COVC18952 | OM738224 | ILRI_COVM01204 | OM738254 |
| ILRI_COVC19669 | OM738225 | ILRI_COVM01221 | OM738255 |
| ILRI_COVC19670 | OM738226 | ILRI_COVM01229 | OM738256 |
| ILRI_COVC19994 | OM738227 | ILRI_COVM01234 | OM738257 |
| ILRI_COVC20064 | OM738228 | ILRI_COVM01279 | OM738258 |
| ILRI_COVC20471 | OM738229 | ILRI_COVM01327 | OM738259 |
| ILRI_COVC20624 | OM738230 | ILRI_COVM01348 | OM738260 |
| ILRI_COVC20783 | OM738231 | ILRI_COVM01396 | OM738261 |
| ILRI_COVC20784 | OM738232 | ILRI_COVM01418 | OM738262 |
| ILRI_COVC20803 | OM738233 | ILRI_COVM01624 | OM738263 |
| ILRI_COVC21359 | OM738234 | ILRI_COVM01772 | OM738264 |
| ILRI_COVC22822 | OM738235 | ILRI_COVM00167 | OM738265 |
| ILRI_COVC23476 | OM738236 | ILRI_COVM00073 | OM738266 |
| ILRI_COVC23642 | OM738237 | ILRI_COVM00174 | OM738267 |
| ILRI_COVC23878 | OM738238 | ILRI_COVM00151 | OM738268 |
| ILRI_COVC23936 | OM738239 | ILRI_COVM00103 | OM738269 |

| | | | |
|---|---|---|---|
| ILRI_COVM00185 | OM738271 | ILRI_COVM00237 | OM738270 |
